# Deep Lipidomic Phenotyping Identifies Ceramide-Centered Lipotoxicity and Depletion of Plasmalogen-Carnitine Pathways in Major Depressive Disorder: Implications for Precision Medicine

**DOI:** 10.64898/2026.02.01.26345328

**Authors:** Yingqian Zhang, Xu Zhang, Mengqi Niu, Yiping Luo, Abbas F. Almulla, Annabel Maes, Jing Li, Bo Zhou, Michael Maes

## Abstract

**Background:** Major depressive disorder (MDD) severely impairs individual health and creates heavy societal burdens. Diagnostic and therapeutic research remains hindered by MDD’s marked heterogeneity and the absence of valid biomarkers. As a neuro-immune, metabolic, and oxidative stress (NIMETOX) disorder, MDD exhibits metabolomic signatures as a final common pathway in the Chinese population.

**Objectives:** To identify lipidomic profile differences between MDD patients and healthy controls and examine associations between lipidomic alterations and clinical phenotypes.

**Methods:** We recruited 125 MDD patients and 40 healthy controls, and measured serum lipidomic profiles using liquid chromatography-mass spectrometry. A rigorously controlled multistage machine learning pipeline with leakage-prevention measures was utilized to examine disparities between MDD and control groups and to predict phenome features.

**Results:** We identified 43 differentially abundant lipids between the MDD and control groups. Subsequent factor analysis clustered the 43 lipids into 3 functional modules, namely the increased ceramide/GM3/LNAPE (CERLNAPE) module, the decreased mitochondrial fatty acid oxidation/acetyl-flux (CARSM) module, and the reduced lysophospholipid/ether-lysolipid (LYSOPE) module. The three lipidomic modules correlated with six previously reported metabolomic functional domains, establishing an integrated metabolomics-lipidomics architecture in MDD. A substantial portion of the variance in the overall severity of depression (74.0%), physiosomatic symptoms (58.5%), suicidal ideation (11.1%), and recurrence of illness (36.6%) was associated with the integrated metabolomics-lipidomics architecture.

**Conclusion:** The MDD lipotype indicates a unified metabolic network linked to the NIMETOX pathophysiology of MDD. Lipidomics provides a robust foundation for subtyping and precision psychiatry. Ceramide, acetyl carnitine, lipotoxicity, and plasmalogens are potential drug targets to treat MDD.

## Introduction

Major Depressive Disorder (MDD) is a widely occurring mood disorder, known for its high incidence, low rates of remission, and increased suicide risk (Malhi & Mann, 2018). Nowadays, increasing evidence shows that MDD is a neuro-immune, metabolic, and oxidative stress (NIMETOX) disorder (Maes *et al*., 2025a; Maes *et al*., 2025b; Maes *et al*., 2025c). Dysfunctional lipid metabolism is one of the key factors linking the NIMETOX pathway and MDD (Jirakran *et al*., 2025a). In this regard, reduced cholesterol esterification contributes to depression susceptibility (Maes *et al*., 1994). Decreased serum high-density lipoprotein cholesterol (HDL-C), apolipoprotein A1 (ApoA1), paraoxonase-1 (PON-1), and lecithin-cholesterol acyltransferase (LCAT) serves as biomarkers for MDD and suicidal behaviors, reflecting impaired reverse cholesterol transport (RCT) (Maes *et al*., 1997; Almulla *et al*., 2023). Notably, ApoA1 and HDL are robust lipid biomarkers that act as protective factors in the diagnosis of MDD(Chen *et al*., 2025). Recently, a study reported that MDD is associated with a specific lipid fingerprint, primarily driven by the acute phase response(Chen *et al*., 2025). The co-occurrence of MDD and metabolic syndromes (MetS) reflects a complex interplay among metabolic dysfunction, atherogenic process, and immune alterations, contributing to greater depression severity(Jirakran *et al*., 2025b).

In MDD, studies conducted at multiple molecular levels reveal that depression is based on a complex array of interconnected metabolic pathways(Fries *et al*., 2022; Maes *et al*., 2025d). Specifically, lipotoxicity refers to the harmful effects of excessive fat accumulation in tissues not designed for fat storage. In the brain, lipotoxicity increases the risk of neurodegenerative diseases, neuroinflammation, microglial dysfunction, and impaired long-term synaptic potentiation(Liu *et al*., 2025). Phospholipid remodeling (mainly via the Lands’ cycle) dynamically regulates the fatty acyl chain composition of phospholipids, determining the biophysical properties of biological membranes, thereby supporting core cellular processes such as signal transduction, vesicular trafficking, and molecular transport. It is critical for maintaining systemic lipid homeostasis and organ function(Wang & Tontonoz, 2019). The fatty acid storage and redistribution module points to a compromised buffering of saturated fatty acids, akin to “overflow”, promoting their transformation into bioactive and oxidized lipids that increase lipotoxic stress(Carli *et al*., 2024).

Ether lipids, especially plasmalogen, are prevalent in the nervous system and play a role in structuring neuronal membranes and the myelin sheath. Abnormal levels of plasmalogens have been described in a variety of neurological diseases(Dorninger *et al*., 2020). Mitochondrial dysfunction is detected in multiple brain regions in MDD patients. The buildup of dysfunctional mitochondria accelerates neuronal dysfunction, while damaged mitochondria further exacerbate changes in the brain environment, fostering neuroinflammation and energy exhaustion, thereby worsening the progression of depression(Song *et al*., 2023). A retinoid detoxification and antioxidant deficit module suggests weakened transcriptional and redox support for neuronal maintenance, making limbic circuits more vulnerable to inflammation and stress(Shmarakov, 2015; Hossain *et al*., 2024). These metabolic pathways are key drivers of dysfunction under pathophysiological conditions, including reactive oxidative stress (ROS), inflammation, α-syn accumulation, and myelin debris(Vanherle *et al*., 2025).

Research on lipidomics in MDD has been extensively conducted across various nations. In the Dutch population, the comprehensive profile of depression revealed changes in lipid metabolism, characterized by a decrease in long-chain fatty acids and an increase in lysophospholipids(Jansen *et al*., 2024). Tkachev et al. observed a strong relationship between lipid levels and the severity of depression symptoms reported by individuals, and a lipid-based model can be formulated to effectively distinguish between MDD and non-depressed individuals in the Moscow population(Tkachev *et al*., 2024). In China, Wang et al. identified dysregulation of membrane lipid homeostasis in depressed adolescents compared to healthy controls, including elevated cholesterol, sphingomyelins, and ceramides, along with lipid oxidation, reduced ether lipids, and reduced polyunsaturated fatty acids (PUFAs) and membrane fluidity. Additionally, lipid damage correlated with depressive symptoms and cognition in Chinese patients(Wang *et al*., 2025).

We have recently determined that the depression phenome consists of multiple subdomains, such as melancholia, anxiety, chronic fatigue, autonomic, vegetative symptoms and suicidal behaviors(Niu *et al*., 2025). To evaluate these phenomes, newly generated phenome scores derived from machine learning should be applied (Niu *et al*., 2025). These factors are as follows: 1) a generalized factor extracted from the symptom domain of the current index episode, referred to as overall severity of depression (OSOD); 2) a single group factor encompassing all psychosomatic domains (designated psychosomatic symptoms); 3) current suicidal behaviors; and 4) the recurrence of illness (ROI) index, which is based on the lifetime number of episodes and suicidal behaviors(Niu *et al*., 2025). However, no study has defined the lipotypes of MDD or its related characteristics mentioned above. Comprehensive lipidomics analysis of well-characterized MDD cohorts is critical for identifying distinct lipidomic signatures, which clarify how immune activation and lipidomic dysfunctions modulate neuro-affective toxicity.

Therefore, the present study aims to: 1) Identify alterations in the lipidomic profile of patients with MDD compared with healthy controls; 2) Verify correlations between lipidomic features and MDD phenome, including OSOD, physiosomatic symptoms, SI, and ROI; 3) Explore associations between lipidomic profiles and 6 metabolic modules in MDD, as determined via deep metabolomic screening.

## Methods

### Participants

A total of 165 individuals were recruited for this study, comprising 125 MDD inpatients and 40 healthy controls (HCs). This research employed a case-control cross-sectional design conducted at the International NIMETOX Center, Mental Health Center of Sichuan Provincial People’s Hospital in Chengdu, China. The participants’ ages ranged from 18 to 65 years, demonstrating an equitable gender ratio. Inpatients with MDD were diagnosed according to DSM-5 criteria, were in an acute phase of illness, and demonstrated a Hamilton Depression Rating Scale 21 (HAMD-21) score greater than 18(Niu *et al*., 2025). Hospital staff, their relatives, and patients’ friends formed the control group, matched to the case group by age, gender, education, and body mass index.

Participants in the control group were excluded if they had a diagnosis of MDD, dysthymia, DSM-4 anxiety disorders, or a familial history of mood disorders, substance use disorders (except nicotine dependence), or suicide. The exclusion criteria for this investigation comprised: 1) diagnosis of various significant psychiatric disorders including schizophrenia, bipolar disorder, eating disorders, substance use disorders (excluding nicotine dependence), autism spectrum disorder, psycho-organic disorders, and schizoaffective disorder; 2) neurological conditions such as stroke, multiple sclerosis, epilepsy, brain tumors, Alzheimer’s and Parkinson’s disease; 3) significant allergic response occurring in the preceding month; 4) having undergone an infection within the past three months; 5) systemic disorders include type 1 diabetes, rheumatoid arthritis, chronic obstructive pulmonary disease, inflammatory bowel disease, systemic lupus erythematosus, psoriasis, or cancer; 6) current administration of immunosuppressants, corticosteroids, or other medicines that alter the immune system; 7) being pregnant or lactating; 8) surgery performed within the past three months; 9) antisocial or borderline personality disorder and developmental difficulties; 10) administration of therapeutic dose of antioxidants or Omega-3 supplements in the past three months; and 11) regular intake of analgesics.

All participants or their legal representatives provided written informed consent. The Ethics Committee of Sichuan Provincial People’s Hospital approved the study “Ethics (Research) 2024-203”.

### Clinical Assessment

A trained research physician conducted a semi-structured interview to collect demographic data, medical history, psychological history, and family background. Psychiatric diagnoses were validated through the Mini International Neuropsychiatric Interview (M.I.N.I.), which evaluates various psychiatric disorders both past and present. The assessor handed out various questionnaires to all participants on the same day to evaluate different rating scales measuring the intensity of somatic-psychosomatic symptoms, anxiety, and depression, as previously described (Niu *et al*., 2025).

To assess the severity of MDD, the Hamilton Depression Scale (HAMD)-21 was employed; anxiety severity was measured with the Hamilton Anxiety Scale (HAMA). Self-reported depression levels were measured with the Beck Depression Inventory, and state anxiety was evaluated using the State-Trait Anxiety Inventory state subscale (Hamilton, 1959; 1960; Spielberger *et al*., 1983; Beck *et al*., 1996). The Somatic Symptom Scale-8 (SSS-8) evaluated how severe somatic symptoms were for individuals during the past week (Gierk *et al*., 2014). The FibroFatigue Scale (FFS), consisting of 12 items, was used to assess the severity of symptoms associated with chronic fatigue syndrome (CFS) (Zachrisson *et al*., 2002; Gierk *et al*., 2014). Both lifetime and current suicidal attempts (SA) and suicidal ideation (SI) were measured using the Columbia Suicide Severity Rating Scale (C-SSRS) (Posner *et al*., 2011). As mentioned before, these scales were used to determine OSOD, physiosomatic symptoms, and current suicidal ideation scores (Niu *et al*., 2025). In this study, two items from the C-SSRS (number of lifetime suicidal attempts and ideation preceding the index episode) were used to evaluate the recurrence of illness (ROI) index, which is calculated as a composite score in z units by adding the z-scores for the total number of depressive episodes, suicidal attempts, and suicidal ideation experienced over a lifetime(Maes *et al*., 2024a).

In the present study, the components of MetS were analyzed, covering height, weight, waist circumference (WC), and body mass index (BMI). BMI was calculated by dividing body weight (in kilograms) by height (in meters) squared. Measurements of waist circumference were conducted halfway between the iliac crest and the lower rib. The definition of MetS was established in the 2009 joint statement from the American Heart Association and the National Heart, Lung, and Blood Institute (Alberti *et al*., 2009). MetS was diagnosed when three or more of the following five criteria were satisfied: (1) male waist circumference ≥90 cm or female waist circumference ≥80 cm; (2) triglycerides ≥150 mg/dL; (3) male HDL cholesterol levels below 40 mg/dL or female HDL cholesterol levels below 50 mg/dL; (4) systolic blood pressure of 130 mm Hg or higher, diastolic blood pressure of 85 mm Hg or higher, or the use of antihypertensive drugs; (5) fasting glucose levels of 100 mg/dL or higher or a diagnosis of diabetes mellitus.

### Assays

Blood samples were obtained between 06:30 and 08:00 hours, with 30 mL of fasting venous blood extracted using a reusable syringe and subsequently transferred into serum tubes. Following centrifugation at 3500 rpm, the serum was collected, aliquoted into Eppendorf tubes, and stored at -80°C for later analysis.

#### Metabolite Extraction

200 μL samples were mixed with 400 μL cold MTBE and 80 μL ice-cold methanol, centrifuged at 3000 rpm for 15 min; 200 μL supernatant was freeze-dried, reconstituted with 200 μL dichloromethane/methanol (1:1, v/v), centrifuged again (3000 rpm, 15 min), and the supernatant was subjected to UPLC-HRMS. QC samples were prepared by pooling equal volumes of all sample supernatants.

#### Chromatography

Separation was performed on an ACQUITY UPLC CSH C18 column (100 mm × 2.1 mm, 1.7 µm, Waters). Mobile phase A (acetonitrile/water, 6:4 + 10 mmol/L ammonium formate + 0.1% formic acid) and phase B (isopropanol/acetonitrile, 9:1 + 10 mmol/L ammonium formate + 0.1% formic acid) were used with gradient elution (30%–100% B in 8.6 min, re-equilibrated to 30% B), flow rate 0.3 mL/min, injection volume 4 µL, column temperature 40℃.

#### Mass Spectrometry

Lipids were detected using a Triple TOF 6600 (AB SCIEX) high-resolution tandem mass spectrometer in positive/negative ESI modes (ESI temp 500℃, voltages +5000 V/-4500 V; curtain gas 30 psi, Gas 1/Gas 2 60 psi). Data were acquired in full scan (50–2000 Da, 170 ms) and IDA modes (top 12 ions with intensity >100, 25–1200 Da, 30 ms, dynamic exclusion 4 s). See the detailed methods in electronical supplementary files (ESF).

### Statistics

#### Classical statistical tests

Contingency table analysis was employed to investigate relationships among categorical variables, using Chi-square testing. An analysis of variance was used to assess differences across the study groups for continuous variables, including demographic and clinical data.

The sample size estimate was derived from the findings of Maes et al. (different papers), which indicated that a significant portion of the variance in OSOD, i.e., more than 25% could be anticipated using multivariable regression analysis, including metabolic or NIMETOX variables. Power calculation for the primary statistical analysis of this study, specifically multiple regression analyses examining the predictors of the phenome scores, was conducted using G*Power 3.1.9.4 software. The analysis utilized an effect size of 0.33 (approximately 25% of the phenome explained), an alpha level of 0.05 (two-tailed), a power of 0.8, and a maximum of seven covariates. The power analysis indicated a minimum necessary sample size of 51 based on the determined effect size. The sample size was increased to include a testing and a holdout sample.

#### LC-MS raw data processing

MS data preprocessing (peak picking, grouping, retention time correction, secondary grouping, isotope and adduct annotation) was performed with XCMS. Raw LC-MS data were converted to mzXML and analyzed via XCMS, CAMERA and metaX in R. Ions were defined by RT-m/z pairs, generating a three-dimensional matrix of peak indices, samples and ion intensities. Metabolites were annotated against KEGG and HMDB databases using exact m/z values (mass error <10 ppm), with chemical formulas validated by isotopic distribution; internal fragment-spectrum library was also used for identification confirmation.

#### Machine learning for biomarker discovery

Biomarker discovery was performed utilizing a supervised multistage pipeline that included principal component analysis (PCA), linear discriminant analysis (LDA), partial least squares discriminant analysis (PLS-DA), orthogonal PLS-DA (OPLS-DA), PLS-regression, support vector machines (SVM), and stringent sample splitting techniques to prevent model overfitting or circularity. Statistical analysis was predominantly performed utilizing R (v4.0), Statistica 14.0, SPSS 30.0, and the Unscrambler. Cluster heatmaps were produced using the R package heatmap. The volcano map, heatmaps, PCA, PLS-DA, and OPLS-DA figures were generated using the OmicStudio tool at https://www.omicstudio.cn/tool.

We performed PCA with varimax rotation to identify latent constructs in lipidomic data, using KMO > 0.7 for sampling adequacy and retaining principal components explaining >50% variance with factor loadings >0.6.

Given the compositional nature of lipidomic data, we applied centered log-ratio (CLR) transformation in R. Multivariate differences between MDD and HC were analyzed using full (n=1095) and curated endogenous lipid datasets (n = 157) in R (v4.0). Data processing included missing value filtering, KNN imputation, and PQN normalization. Heatmaps were generated with pheatmap; PCA, differential analysis, and VIP scoring used ropls; correlation analysis employed Pearson’s coefficient. Differential metabolites were defined by t-test *p <* 0.05, fold change >1.2, and PLS-DA VIP ≥1. PLS-DA models were evaluated via R^2^Y, Q^2^, and CV-ANOVA, with VIP identifying key variables. This approach was solely for exploratory multivariate analysis, not biomarker identification, prediction, or validation.

#### Testing procedures

The 43 differential lipids and 3 derived module scores were subsequently employed in the real model evaluation phase within the testing population. To assess the replicability of categorization, we first applied a z-score transformation to the test set data and subsequently recalculated the composite scores within this limited sample. The trained models, employing either 43 lipids or 3 (+6 metabolomics) modules, were later analyzed using LDAs applied exclusively to the testing sample, applying leave-one-out (LOO) cross-validation to evaluate out-of-sample prediction stability. The technique ensured a strict distinction between variable selection and model evaluation, hence guaranteeing unbiased biomarker discovery and preventing data leakage. See the detailed methods in electronical supplementary files (ESF).

#### Regression methods

We predicted phenome scores (OSOD, physiosomatic, current SI, and ROI) in the aggregated training and testing sets using PLS regression. The model’s accuracy was evaluated using R^2^Y and Q^2^ values for all latent vectors. We produced score contribution profiles or case-level contributions for each subject. The method shows how lipids increase a subject’s phenome scores. This provides each forecast with a molecular basis and identifies which lipid aberrations drive sickness severity in each individual, an approach increasingly needed for precision psychiatry.

We used manual multiple regression and automated regression to determine lipid scores, which predict phenome features. A ridge regression analysis was performed in Statistica (Version 14, Windows) with λ = 0.1 and a tolerance level of 0.4. We also used forward stepwise automatic linear modeling analyses using an overfit criterion for variable inclusion and exclusion and a maximum effects threshold of 5 (SPSS version 30, Windows). We performed a “best subset” analysis to prevent overfitting, using 6 metabolomic functional domains and 3 lipidomic modules from the SPSS regression. After these assessments, we conducted a manual regression analysis to analyze model statistics, including F statistic, degrees of freedom, p-value, and R^2^ (total variance). We also examined each predictor’s standardized beta coefficients, t-statistics, and exact p-value. The residual distributions and P-P plots were used to test multivariate normality in the final models. To detect collinearity of multicollinearity, the variance inflation factor and tolerance were examined. The White and modified Breusch-Pagan tests assessed heteroskedasticity and homoscedasticity. Partial regression analysis of lipidomic phenome data was also undertaken. The regression analysis was done using IBM SPSS, Windows 30, and Statistica 14.0. The significance level for all statistical analyses was 0.05 using two-tailed testing.

## Results

### Demographic and clinical data

The Electronic Supplementary File (ESF) Table 1 indicates no significant differences in age, sex distribution, MetS, BMI, and education between MDD inpatients and controls. Patients with MDD had elevated scores in psychosomatic symptoms, suicide ideation, ROI, and OSOD compared to the control group.

**Table 1.** Results of factor analysis conducted on lipids.

| **Variables** | **PC1** |
| --- | --- |
| LNAPE 18:1/N 18:0 | 0.624 |
| Cer 18:1;2O/24:2 | 0.657 |
| Cer 18:1;2O/22:1 | 0.626 |
| Cer 18:1;2O/24:1 | 0.742 |
| GM3 36:1;2O | 0.725 |
| Cer 18:1;2O/23:0 | 0.671 |
| GM3 42:1;2O | 0.730 |
| PE O-20:3_22:6 | 0.791 |
| Cer 18:1;2O/22:0 | 0.710 |
| Cer 18:1;2O/24:0 | 0.653 |
| Cer 18:1;2O/16:0 | 0.827 |
| LNAPE 20:4/N-17:0 | 0.666 |
| GM3 36:2;2O | 0.709 |
| Cer 30:1;3O/12:0;(2OH) | 0.824 |
| LNAPE 16:1/N-18:0 | 0.735 |
| PE(16:0/22:6(4Z,7Z,10Z,13Z,16Z,19Z)) | 0.653 |
| Cer(d18:1/24:1) | 0.860 |
| Cer(d18:1/25:0) | 0.725 |
| Cer 18:2;2O/23:0 | 0.818 |
| Cer 24:3;2O/18:2;(2OH) | 0.802 |
| PE(P-16:0/22:6(4Z,7Z,10Z,13Z,16Z,19Z)) | 0.770 |
| GM3 34:1;2O | 0.719 |
| Cer(d18:1/20:0) | 0.863 |
| Cer 27:3;2O/16:2;(2OH) | 0.752 |
| Cer 14:0;2O/29:1;(2OH) | 0.749 |
| Cer 26:3;2O/16:2;(2OH) | 0.691 |
| Cer 26:3;2O/18:2;(2OH) | 0.827 |
| **KMO** | 0.905 |
| **Bartlett's test of sphericity$\chi^2$** | 11186.48 |
| **df** | 351 |
| ***p*** | < 0.001 |
| **Designated as** | **CERLNAPE module** |

### Lipidomics in MDD versus controls

The initial metabolomics dataset had 1,095 identified lipids. **Figure 1** illustrates a volcano plot using log2 fold change, demonstrating a notable pattern of differential regulation in MDD versus controls. Ninety-two lipids were markedly increased, while 65 were substantially decreased. The remaining metabolites showed no significant alteration, creating a neutral backdrop against which to contrast the altered lipids.

**Figure 1.**
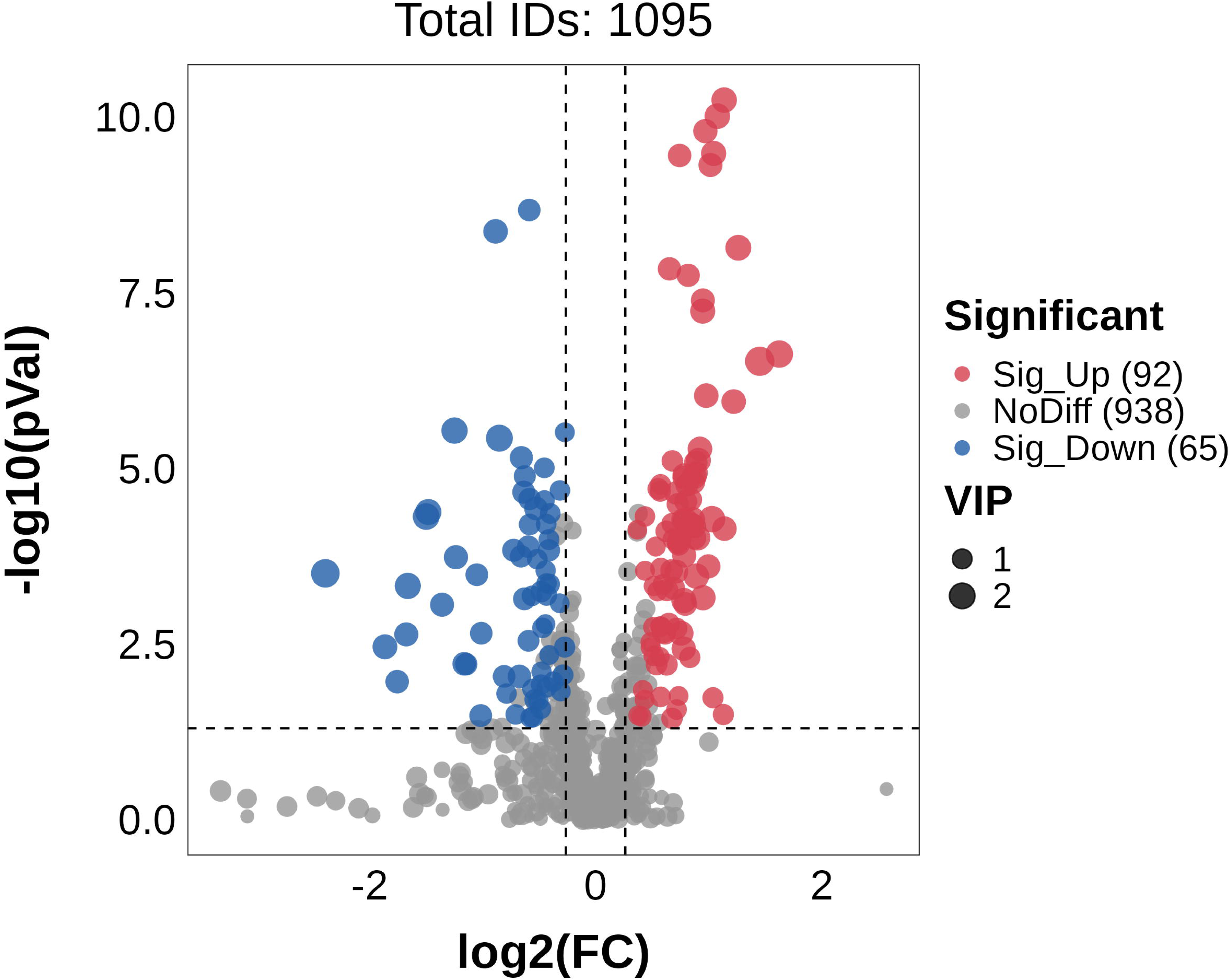
Volcano plot, with log2 fold change, of 1095 lipids in patients with major depressive disorder and healthy controls.

Consequently, we compiled a curated lipid file, omitting exogenous, drug-related, artifact-derived, or low-confidence metabolites, and retaining only those with VIP > 1.2 and FDR < 0.05. The study of the selected data set, comprising 43 high-confidence lipids, revealed that the multivariate structure was perfectly consistent with the PLS-DA analysis of the original data set of 1095 lipids. **Figure 2** illustrates the outcomes of a heatmap depicting the distinctions between the two classes, as well as the results of PCA, PLS-DA, and OPLS-DA analyses. **Figure 2A** presents a heatmap of the 43 selected lipids, showing that patients with MDD display distinct, block-like patterns of upregulated and downregulated lipids across the matrix. Nonetheless, certain patients exhibit both upregulated and downregulated lipid profiles akin to those of controls, whereas a minority of controls display upregulated lipid profiles similar to those of patients. ESF, Figure 1 illustrates the outcomes of the ROC analysis conducted on the entire dataset and the five top upregulated lipids. **Figure 2B** demonstrates the score plot of PC1 against PC2, revealing distinct patterns between the two groups in the two-dimensional space, despite some overlap between the classes. PC1 accounts for 46.34% of the variance and serves as the primary distinguishing axis.

**Figure 2.**
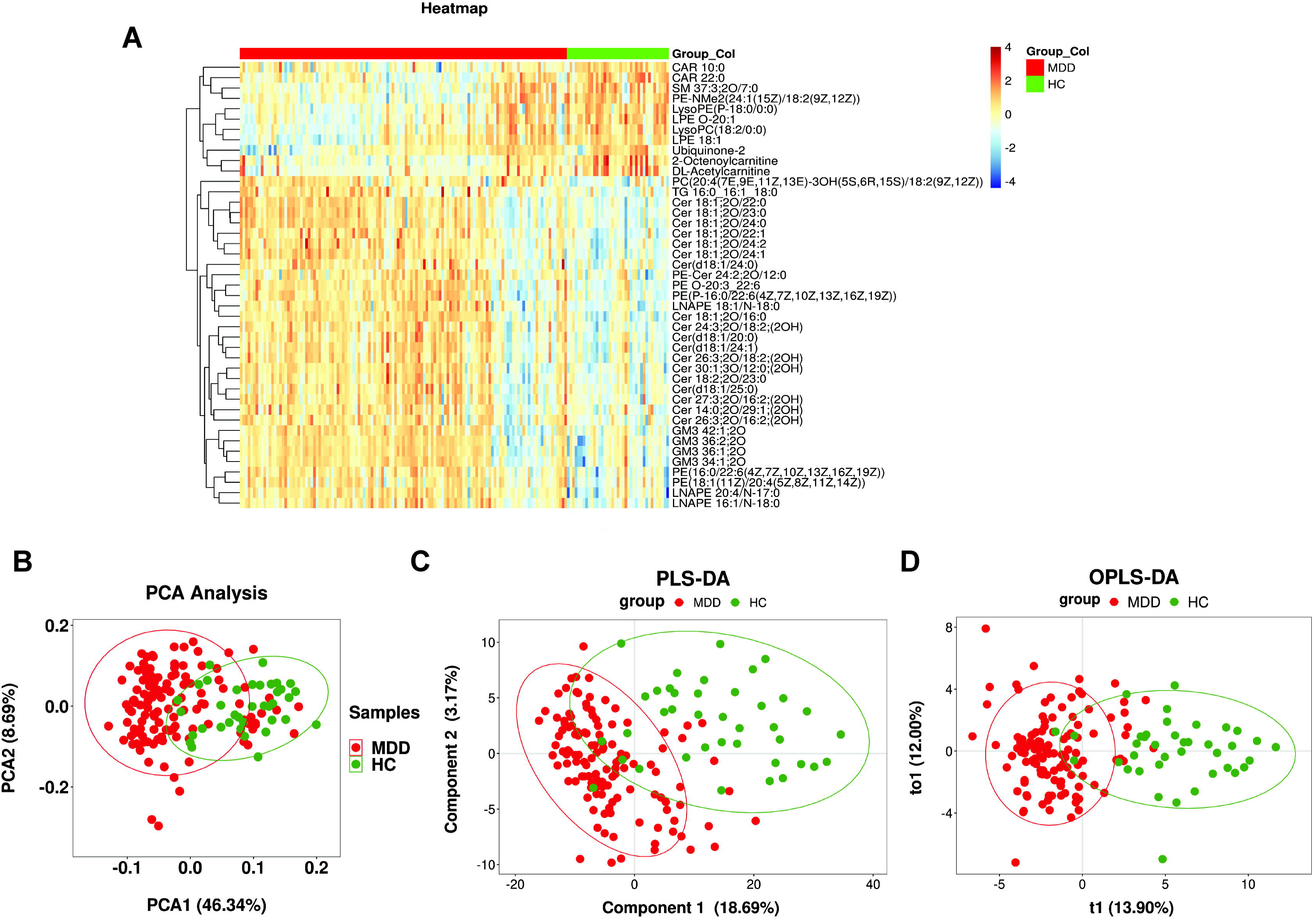
Differential lipids between the MDD and HC groups. **Figure 2A:** Heatmap based on 43 differential lipids **Figure 2B:** Principal component analysis (PCA) plot **Figure 2C:** Partial least squares discriminant analysis (PLS-DA) model **Figure 2D:** Orthogonal PLS-DA (OPLS-DA) plots. PCA, PLS-DA, and OPLS-DA show a clear separation trend between MDD and HC, even though there is some overlap.

**Figure 2C** illustrates the PLS-DA model, revealing discrete clusters, but a limited number of patients and controls are assigned to the alternative class. The PLS-DA model depicted in Figure 2C comprises two significant components and achieves satisfactory performance metrics, with a cumulative Q² of 0.728, a cumulative R²Y of 0.848, and an R²Y of 0.523, indicating substantial discrimination and considerable explained variation in group categorization and predictors. Permutation testing confirmed that the observed model performance was not due to random variation, as all permuted R² and Q² values were significantly inferior to those of the empirical model. **Figure 2D** shows the results of OPLS-DA, with a similar separation between the groups as PLS-DA. The first predictive component (t1) accounted for 13.90% of the variance associated with class separation, and the second orthogonal component (to1) accounted for 12.00% of the variance not connected to class membership. The OPLS-DA model exhibited strong explanatory and predictive performance (R^2^Y = 65.5%, Q^2^ = 47.6%) and statistical significance (p = 0.005), indicating a stable, non-overfitted discriminant model. Misclassifications in the two-dimensional plots were confined to a restricted number of instances.

**Figure 3** presents the importance plot derived from PLS-DA, highlighting that Cer 18:1; 2O/24:1, Cer 18:1; 2O/24:0 (belonging to the CERLNAPE module), and PE (18:1(11Z)/20:4(5Z,8Z,11Z,14Z)) were among the most distinguishing lipids.

**Figure 3.**
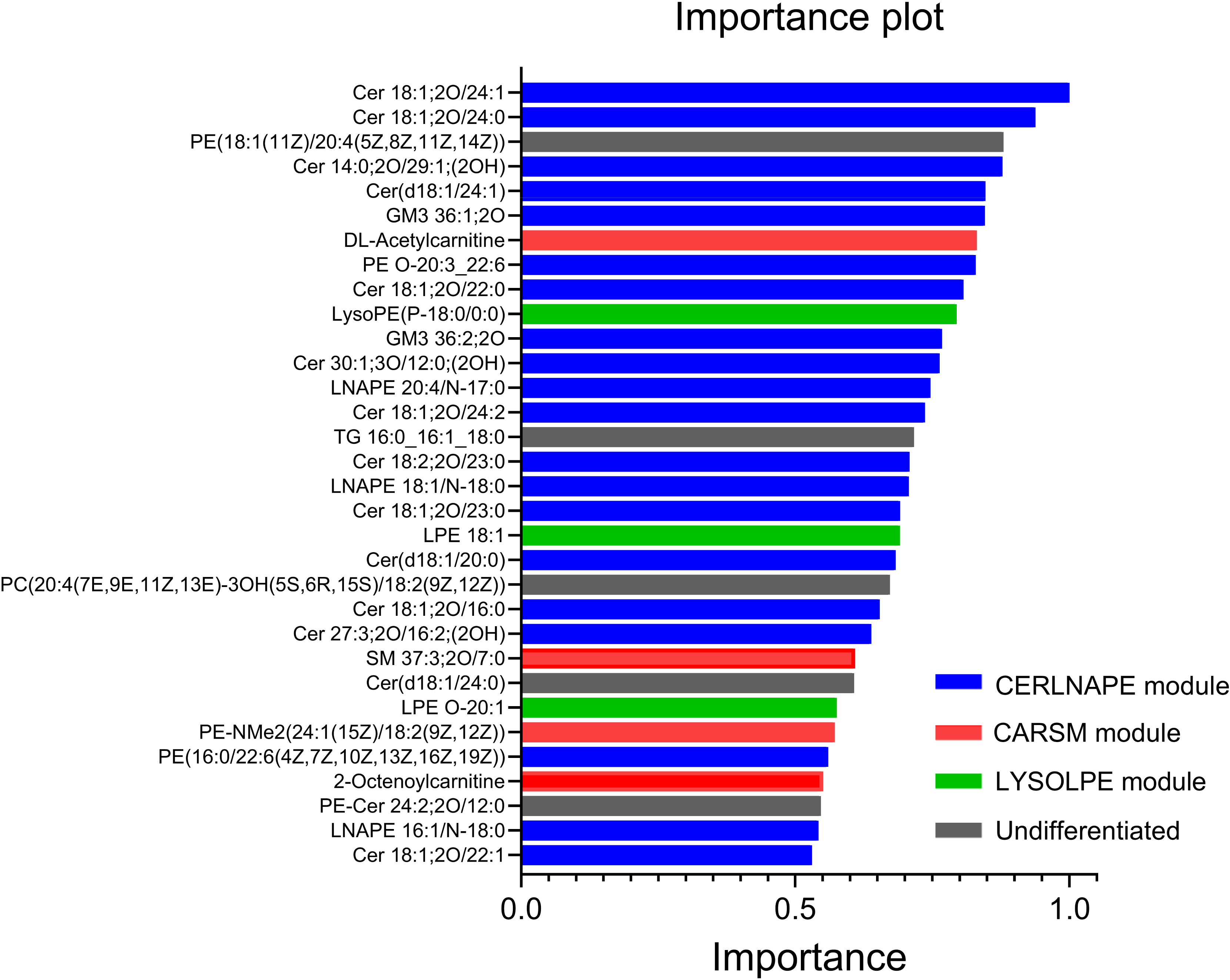
The importance plot of the differential lipids using the PLS-DA model

**Figure 4A** presents a heatmap illustrating the relationships between clinical data and lipid modules. Age had a substantial association with 26 lipids, while sex (shown as point-biserial correlations) had an association with 10 lipids. BMI showed relationships with five lipids, while MetS demonstrated point-biserial correlations with four lipids. Both BMI and MetS significantly influenced Cer 14:0;20/29:1;(2OH).

**Figure 4.**
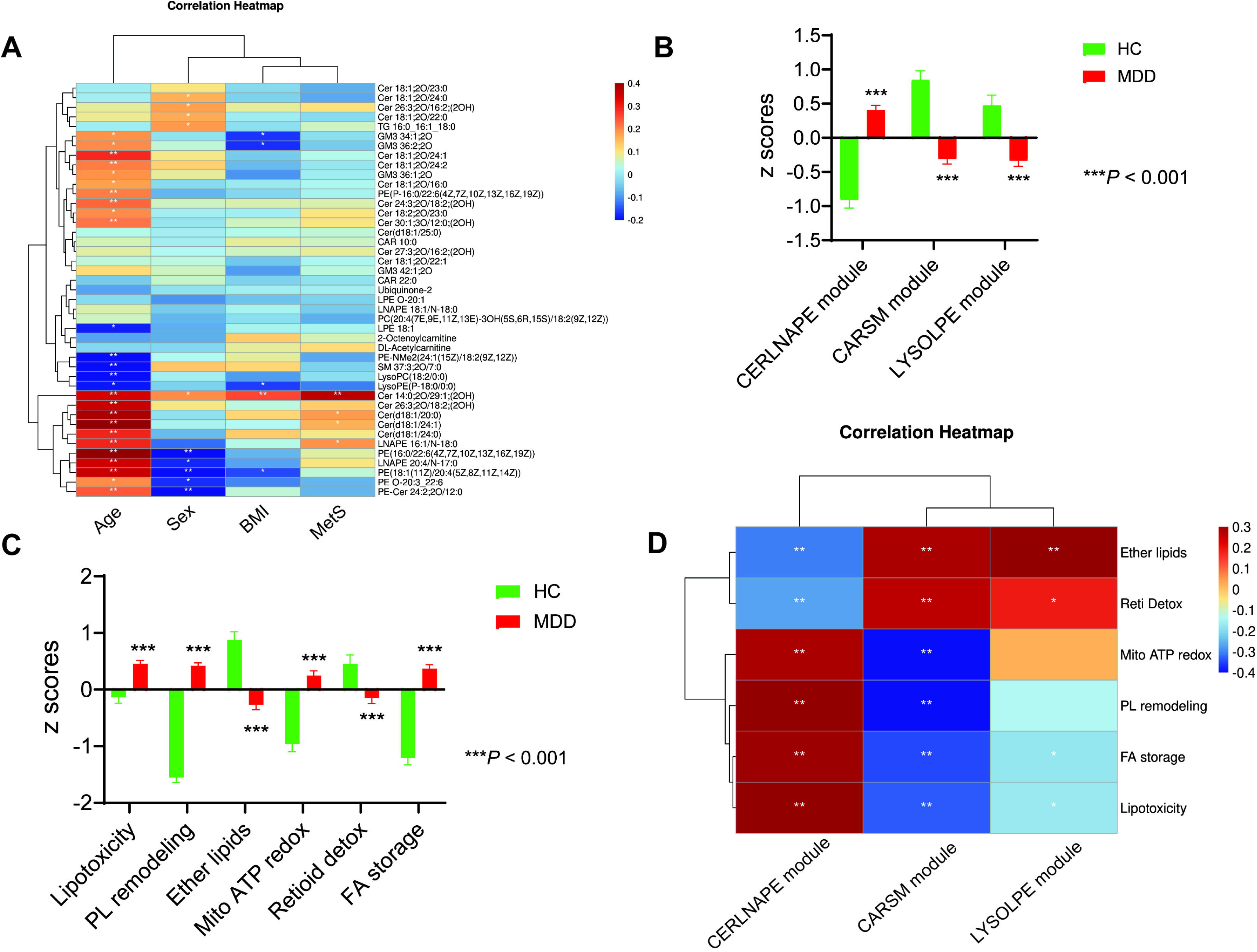
Correlations of basic information and differential lipids and changes of lipids and functional domains in the two groups. **Figure 4A:** Correlation heatmap on 43 differential lipids and basic information (age, sex, BMI, and Mets). **Figure 4B:** Changes in 3 lipidomic modules between MDD and HC groups. **Figure 4C:** Changes in 6 metabolomic functional domains between MDD and HC groups. **Figure 4D:** Correlation heatmap on 3 lipidomic modules and 6 metabolomic functional domains.

### Lipid modules built using PCA

**Table 1** presents the outcomes of PCA. A generalized factor could be derived from 27 lipids. The initial PC accounted for 54.91% of the variance (KMO = 0.905, Bartlett’s test of sphericity χ^2^ = 11186.48, df = 351, *p* < 0.001). All lipids, including ceramides, LNAPE, and GM lipids, were positively associated with this principal component, which was subsequently designated as the upregulated CERLNAPE module (from ceramides and LNAPE). We could not extract a single PC from the remaining variables; however, varimax rotation indicated that two rotated principal components may be obtained (**Table 2**). The first component accounted for 31.83% of the variance, while the second accounted for 29.68%, resulting in a cumulative total of 61.52% (KMO = 0.749, Bartlett’s test of sphericity χ^2^ = 1233.78, df = 55, *p* < 0.001). The initial varimax-rotated component exhibited significant loading on SM 37:3;2O/7:0, CAR 22:0, CAR 10:0, 2-Octenoylcarnitine, DL-Acetylcarnitine, and PE-NMe2(24:1(15Z)/18:2(9Z,12Z)) and was, therefore, designated as the CARSM module (from carnitines and SM). The second varimax-rotated factor exhibited significant loading on LysoPC(18:2/0:0), LysoPE(P-18:0/0:0), LPE 18:1, and LPE O-20:1, and was designated as the LYSOLPE module (from lyso lipids and LPE).

**Table 2.** Results of varimax rotation conducted on lipids.

| Variables | Varimax rotated PCs |  |
| --- | --- | --- |
|  | Loadings PC1 | Loadings PC2 |
| SM 37:3;2O/7:0 | **0.706** | 0.169 |
| LysoPC(18:2/0:0) | 0.169 | **0.920** |
| LysoPE(P-18:0/0:0) | 0.204 | **0.886** |
| LPE 18:1 | 0.051 | **0.880** |
| LPE O-20:1 | 0.205 | **0.861** |
| CAR 22:0 | **0.716** | 0.203 |
| CAR 10:0 | **0.659** | -0.037 |
| Ubiquinone-2 | 0.423 | 0.356 |
| 2-Octenoylcarnitine | **0.860** | 0.136 |
| DL-Acetylcarnitine | **0.667** | 0.163 |
| PE-NMe2(24:1(15Z)/18:2(9Z, 12Z)) | **0.704** | 0.171 |
| **KMO** | 0.749 |  |
| **Bartlett's test of sphericity $\chi^2$** | 1233.78 |  |
| **df** | 55 |  |
| ***p*** | < 0.001 |  |
| **Total variance explained** | 61.52% |  |
| **Variance explained** | 31.83% | 29.68% |
| **Designated as** | **CARSM module** | **LYSOLPE module** |

Therefore, we utilized the PC scores from these three modules in univariate, multivariate, and ML analyses. The CERLNAPE score showed a strong inverse correlation with CARSM (r = -0.710, *p* < 0.001) and LYSOLPE (r = -0.511, *p* < 0.001). **Figure 4B** represents the PC scores for both control subjects and patients with MDD. Factorial ANCOVA was used with MDD and MetS as fixed factors, and age, sex, smoking, and BMI as covariates. Given the absence of substantial effects of the last three covariates and MetS (even without FDR *p-*value correction), we present the GLM analysis with age as the sole covariate. All three module scores exhibited significant differences between the two groups, with CERLNAPE scores being elevated in MDD, while the CARSM and LYSOLPE component scores were diminished in MDD compared with controls. Age had a significant impact on the CERLNAPE score (t = +4.78, *p* < 0.001) and the LYSOLPE score (t = -2.39, *p* = 0.018). Interestingly, MetS demonstrated no effects on any of the three modules, even without *p*-value adjustment. Moreover, the same results regarding the three lipid modules were observed in individuals without MetS. Our findings indicate that 92 patients with MDD were administered antidepressants, 68 were given benzodiazepines, 44 received atypical antipsychotics, and 10 were provided with mood stabilizers. These drugs did not have a significant impact on the three lipid domains, even after FDR *p*-value correction (results of multivariate GLM analysis). Thus, there is no evidence suggesting that the patient’s medication status affected the research outcomes.

### Associations between the 3 lipid modules and 6 metabolic modules

**Figure 4C** presents the six lipid module scores for both MDD and control subjects (multivariate GLM). Age, smoking, MetS, and BMI did not significantly affect these 6 lipid modules (ESF Table 2). Sex was the sole significant variable in this model; hence, we provide the findings of the GLM analysis with sex as the secondary factor. All metabolomic profiles exhibited substantial differences between the two groups. PL remodeling was markedly greater in women compared to men (F = 7.23, df = 1/149, *p* = 0.008), exhibiting a minimal effect size of 4.3%.

Figure 4D illustrates the correlations between the three lipid modules developed in this lipidomics study and the six metabolomics modules established before(Maes *et al*., 2025d; Maes *et al*., 2026). The CERLNAPE and CARSM module scores had significant associations with all metabolomics modules, while the LYSOLPE module demonstrated a positive correlation with ether lipids and Reti Detox, and an unfavorable relationship with lipotoxicity and FA storage modules.

### Associations of the lipids with neuropsychiatric rating scale scores

**Figures 5A and 5B** present heatmaps illustrating the associations between neuropsychiatric ratings and lipids, as well as lipid modules. Significant relationships existed between ROI, OSOD, and psychosomatic symptoms with nearly all lipids, save ubiquinone. The current SI exhibited a significant association with 24 out of 43 lipids. Only 4 significant associations were observed between ACEs and lipids (three upregulated and one downregulated), but sexual abuse exhibited no effect. **Figure 5B** illustrates the correlation matrix among the identical clinical ratings and the three lipid modules. The CERLNAPE module had a substantial positive correlation with OSOD, physiosomatic symptoms, ROI, and present suicidal ideation. The CARSM module had a substantial inverse correlation with these clinical ratings, while the LYSOLPE module demonstrated an association with OSOD and physiosomatic symptoms. No significant relationships were found between the lipid modules and ACEs/sexual abuse.

**Figure 5.**
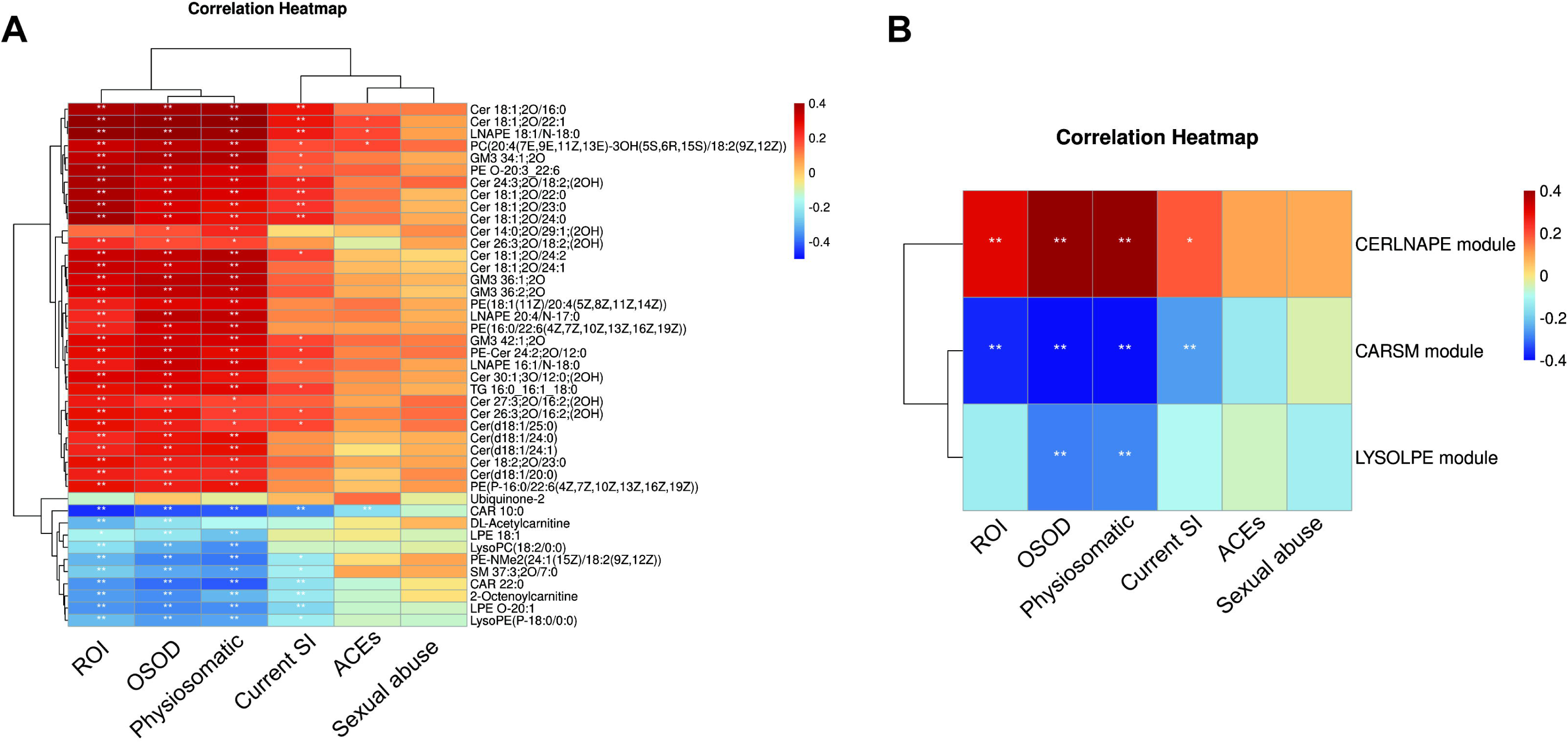
Correlation of differential lipids or lipidomic modules and clinical markers. **Figure 5A**: Correlation heatmap on 43 differential lipids and clinical markers; **Figure 5B**: Correlation heatmap on 3 lipidomic modules and clinical markers.

To assess how the three lipidomics modules (with and without the six metabolomics modules) predicted the clinical data, we conducted multiple regression analysis, treating the clinical data as dependent variables and the nine lipid-metabolomic data as explanatory variables, while allowing for the influences of age, sex, MetS, BMI, smoking, and education. **Table 3** presents the results of various regression studies, with OSOD, psychosomatic symptoms, suicidal ideation, and ROI as dependent variables.

**Table 3.**
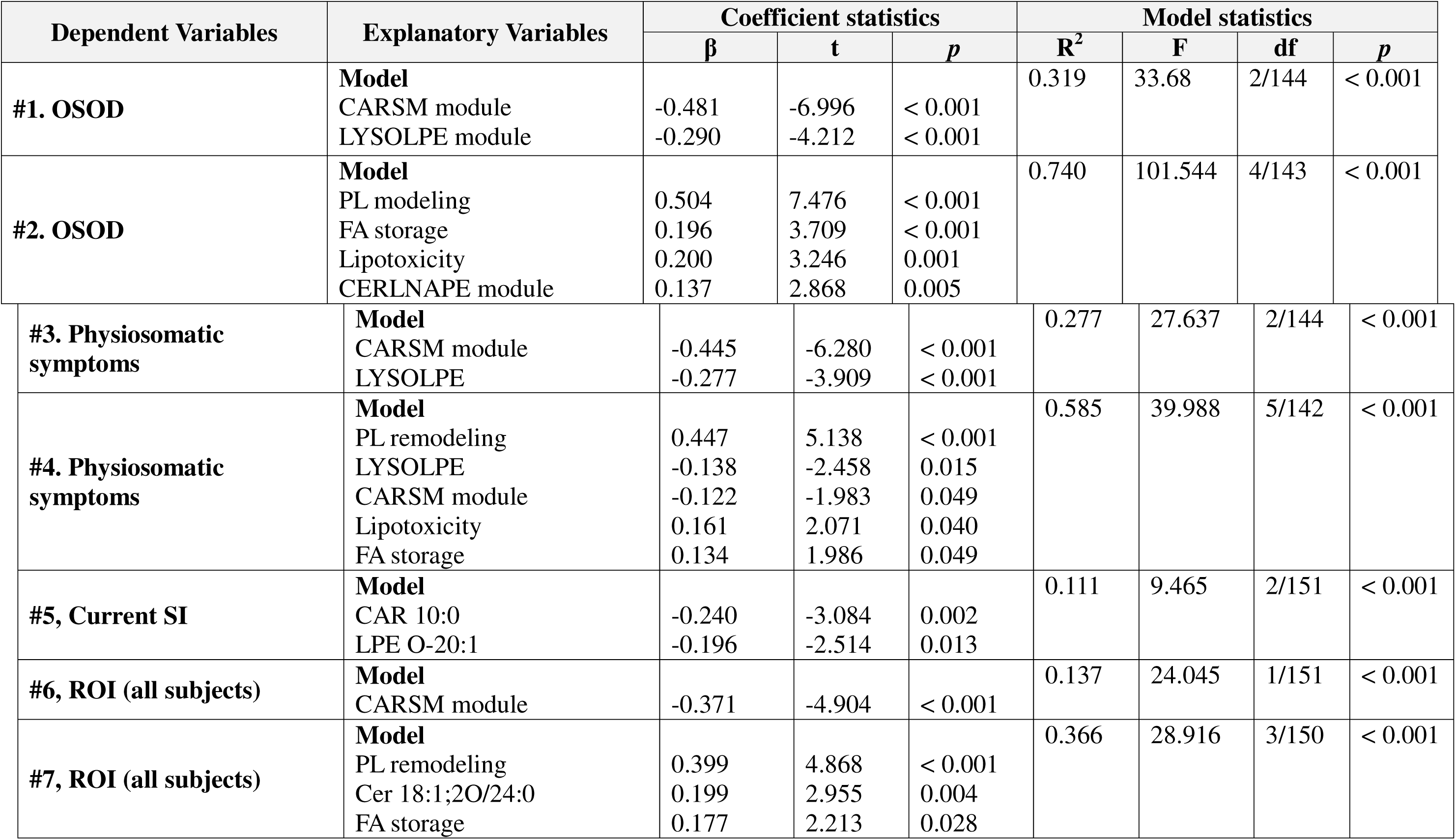
Results of multiple regression analysis with phenome as the dependent variable and lipids as the explanatory variables.

**Table 3**, regression #1 indicates that 31.9% of the variance in OSOD was accounted for by the CARSM and LYSOLPE modules. Upon adding the six metabolomics modules, we determined that 74.0% of the variance in OSOD was accounted for by the regression, including three metabolomic modules in conjunction with the CERLNAPE module, all of which exhibited positive correlations (regression #2).

**Table 3**, regression #3, indicates that 27.7% of the variance in PC physiosomatic was accounted for by the CARSM and LYSOLPE modules, both of which exhibited an inverse relationship. Upon adding the six metabolomics modules, we determined that 58.5% of the variance in PC physiosomatic was elucidated by the regression on three metabolomic modules (both positively correlated) and the CARSM and LYSOLPE modules (both inversely correlated, regression #4).

CARSM scores exhibited a negative correlation with current SI, characterized by a modest effect size (R^2^ = 0.058, F = 9.23, df = 1/151, *p* = 0.003). Consequently, we have investigated the impact of individual lipids on suicidal thoughts. Regression #5 indicates that CAR 10:0 and LPE O-20:1 accounted for 11.1% of the variance in present suicidal ideation. **Table 3**, regression #5 indicates that 13.7% of the variance in ROI was elucidated by the CARSM module (inversely correlated, regression #6). Upon allowing for the effects of the six metabolomics modules, 36.6% of the variance in ROI was elucidated by PL remodeling, FA storage, and Cer 18:1;2O/24:0, all of which exhibited positive correlations (regression #7).

### The personalized approach

**Figure 6** depicts the score contribution profiles for four people, highlighting the contributions from three lipidomic and six metabolomic modules. The latter relies on PLS-DA, which produced two significant components with a cumulative R^2^Y of 0.874, cumulative R^2^X of 0.536, and cumulative Q^2^ of 0.818. Marked heterogeneity was observed between the control group and patients, as well as among the three MDD patients, as illustrated in this figure. **Figure 6A** illustrates a standard control profile characterized by elevated ether lipids, Reti detox, CARSM, and LYSOLPE scores. MDD1, MDD2, and MDD3 exhibit various potential combinations of the nine biomarkers linked to MDD.

**Figure 6:**
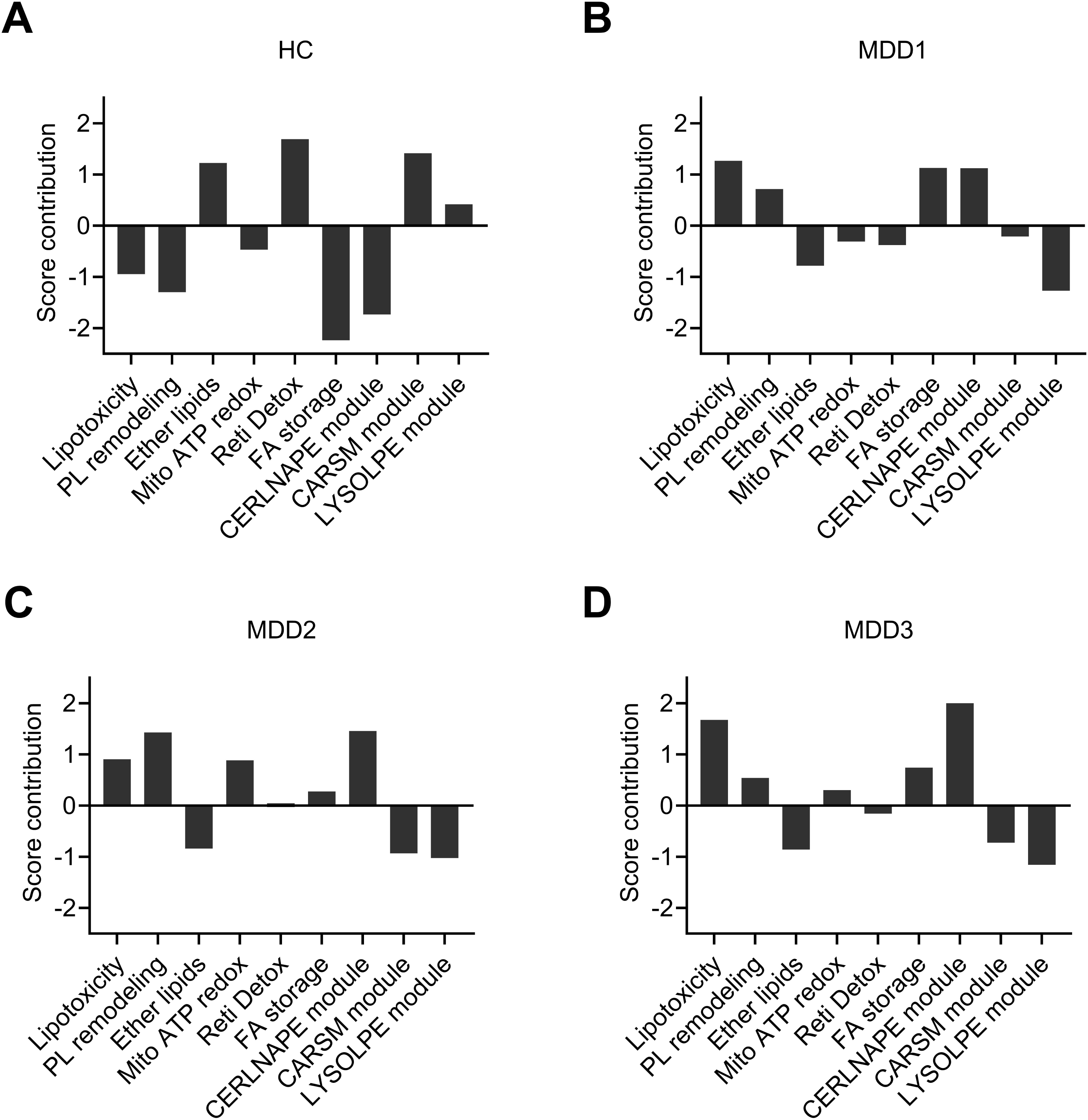
Case-wise score contributions based on 6 metabolomic functional domains and 3 lipidomic modules using PLS regression, showing the personalized profiles based on relative power predicting the severity of MDD in one healthy control and three MDD patients.

## Discussion

### The Three Featured Lipid Modules in MDD

The first significant discovery of this work is the featured lipotypes between MDD and HC. We identified 43 significantly different lipids, which could be clustered into three lipid modules: CERLNAPE, CARSM, and LYSOLPE through factor analysis. The lipidomic characteristics significantly predict the overall severity of depression, physiosomatic symptoms, current suicidal ideation, and the recurrence of illness. The ROI index exhibited a robust correlation with a lipidomic module that may exacerbate the NIMETOX pathway(Maes *et al*., 2025b). Consequently, the lipidomic profiles of MDD and its phenome characteristics correspond with a coherent NIMETOX-associated lipidomic phenotype, which will be further detailed in the following sections.

The significantly altered lipids and modules in our investigation were only weakly influenced by sex, BMI, and MetS, suggesting a lipidomic profile that reflects essential pathophysiological processes in MDD rather than metabolic comorbidities. Furthermore, although no correlations were identified between the lipids and MetS, significant connections were observed with characteristics of MDD. Thus, the lipidomic irregularities in MDD indicate a non-MetS, immune-redox lipid-remodeling phenotype in MDD.

The first lipidomics module, the Ceramide-GM3-LNAPE (CERLNAPE) module, is characterized by increased hydroxylated and very long-chain ceramides, elevated GM3 gangliosides, and increased LNAPE species, with partial enrichment of docosahexaenoic acid (DHA)/eicosapentaenoic acid (EPA)-containing PE and plasmalogen-PE, representing the lipotoxic effector arm of the system. Targeted lipidomics demonstrated pronounced ceramide derangements in MDD(Brunkhorst-Kanaan *et al*., 2019), while mechanistic studies linked ceramide accumulation to depressive-like behavior and showed that ceramide neutralization ameliorates these effects(Schumacher *et al*., 2022).

The second lipidomics module, named CARSM, is a decreased mitochondrial fatty acid oxidation/acetyl-flux module, including reduced CAR 22:0, CAR 10:0, 2-octenoylcarnitine, acetylcarnitine, PE-NMe2, and protective sphingomyelins. The focus is on diminished fatty acid β-oxidation and compromised acetyl-CoA buffering, rather than on lipid overload. According to Nasca et al., there is a deficiency of acetyl-L-carnitine in MDD, which is linked to impaired hippocampal plasticity and the severity of depression, suggesting a bioenergetic-neuroplastic bottleneck(Nasca *et al*., 2018). Large-scale population studies further relate circulating acylcarnitine patterns to depression status and symptom severity, supporting the consistent alteration of carnitine-mediated bioenergetics in MDD(Montanari *et al*., 2025). Complementary lipidomic case-control findings from the Chinese population also highlight dysregulated acylcarnitines and oxidized lipid stress among depressed patients(He *et al*., 2025).

The third lipidomics module, lysophospholipid/ether-lysolipid (LYSOLPE) module, manifested as decreased LysoP(18:2), LysoPE(P-18:0), LPE 18:1, and LPE O-20:1, implying hypodynamic membrane remodeling, concurrent with the loss of plasmalogen-linked intermediates required for rapid membrane repair and resolution signaling(Ge *et al*., 2025). Because ether lipids function as endogenous antioxidants, the loss of these intermediates corresponds to an oxidative environment, which promotes lipid peroxidation and weakens the resilience of synaptic and endothelial membranes(Dean & Lodhi, 2017). Genetic epidemiology links MDD liability to ether-phosphatidylcholine species enriched in omega-6/arachidonate pathways, providing a mechanistic bridge between ether-lipid biology and inflammatory signaling(Knowles *et al*., 2017).

Combined integratively, the architecture is consistent with a self-reinforcing loop: failed lysophospholipid/ether lipid remodeling (LYSOPE module) and bioenergetic/carnitine insufficiency (CARSM module) reduce repair capacity and metabolic flexibility, thereby favoring diversion into ceramide/GM3/LNAPE signaling (CERLNAPE module), which further worsens mitochondrial function and redox stress. Longitudinal community lipidomics study supported this broader pattern of coordinated shifts across sphingolipids and glycerophospholipids in relation to depressive phenotypes(Miao *et al*., 2023).

### Integrated Metabolomics-lipidomics Architecture in MDD

By incorporating six metabolomics modules alongside three lipidomics modules, our data describes a layered metabolic architecture related to MDD. These modules are not merely isolated biochemical abnormalities; they constitute a self-reinforcing network in which impaired repair mechanisms, energy insufficiency, and damaging lipid signaling converge to activate immune responses, oxidative stress, and synaptic dysfunction—key elements of the NIMETOX framework.

The first lipidomics module, CERLNAPE, represents the lipotoxic effector arm of the system. Ceramides and GM3 stabilize rigid membrane microdomains that impair insulin, PI3K– Akt, and neurotrophic signaling while promoting mitochondrial stress and innate immune priming, thereby coupling this increased module back to the two decreased modules(Wilkerson *et al*., 2024).

The second lipidomics module, CARSM, showed decreased acylcarnitine levels, diminished PE-NMe2 abundance, and depletion of protective sphingomyelin, which align closely with the mitochondrial-redox stress and fatty acid overflow modules defined in metabolomic profiling. The metabolic signature denotes mitochondrial hypo-energetics, characterized by impaired fatty acid oxidation flux and compromised acetyl-CoA buffering capacity (Xiang *et al*., 2025). These mitochondrial metabolites participate in the regulation of brain plasticity by modulating histone acetylation, inflammatory pathways, and insulin signaling, and are closely related to the pathological mechanisms of depression(Bigio *et al*., 2024).

The third lipidomics module, LYSOLPE, is defined by low acetylcarnitines, reduced PE-NMe2, and loss of protective sphingomyelin species, and it maps tightly onto the metabolomics mitochondrial-redox stress and fatty acid overflow modules. This pattern signifies mitochondrial hypometabolism, with reduced fatty acid oxidation and impaired acetyl-CoA buffering, rather than lipid overload(Hällqvist *et al*., 2025). Acetyl-L-carnitine deficiency has been demonstrated in MDD and linked to impaired hippocampal plasticity and depressive severity, providing a direct connection between altered carnitine metabolism and neurocircuit dysfunction(Post, 2018; Crivelli *et al*., 2024). This cascade establishes a critical mechanistic link to the metabolomics profiling modules that capture the remodeling of arachidonic/adrenic acid-containing phospholipids and the signaling of arachidonic acid (AA)-derived eicosanoids, all of which align with the core tenets of the inflammatory lipid regulatory framework(Bazinet & Layé, 2014).

Crucially, these modules are not independent. The CARSM and LYSOLPE modules represent capacity deficits, whereas the CERLNAPE module represents the downstream toxic output that feeds back to worsen both repair failure and mitochondrial dysfunction. The metabolomics retinoid detoxification/antioxidant deficit module overlays this architecture, likely diminishing transcriptional and redox resilience, thereby rendering the system more susceptible to inflammatory and oxidative stress, in alignment with overarching mitochondrial and oxidative stress frameworks of depression(Huang & Chen, 2020; van der Spek *et al*., 2023; Bisle *et al*., 2025).

In summary, the integration of metabolomics and lipidomics identifies MDD as a condition characterized by inadequate metabolic adaptability, wherein compromised repair and energy systems facilitate the diversion of lipids into sphingolipid- and ganglioside-mediated inflammatory signaling pathways. This approach harmonizes disparate findings from several investigations and emphasizes modular targets—restoring membrane remodeling capacity, enhancing mitochondrial energetics, and mitigating ceramide/GM3 dominance-for precision-focused therapies.

### Upstream Pathways that may Shape the MDD Lipotypes

#### Inflammation and oxidative stress

The integrated metabolomics-lipidomics profile may effectively induce both inflammation and oxidative/nitrosative stress (O&NS) via an interconnected series of processes. Initially, the reduction of plasmalogen/ether lipids (low ether-lysoPE/LPE) eliminates a significant endogenous membrane antioxidant reservoir; cells lacking plasmalogens exhibit increased susceptibility to ROS(Zoeller *et al*., 1999). Secondly, compromised remodeling combined with redox stress promotes the formation of oxidized phospholipids (OxPL/OxPCs), which can activate innate immune co-receptors (CD14/MD-2/CD36) and influence TLR4 signaling pathways(Serbulea *et al*., 2017). Third, the ceramide/GM3 effector module serves as a direct inflammatory catalyst: ceramide facilitates NLRP3 inflammasome formation and IL-1β secretion in microglia(Scheiblich *et al*., 2017). Concurrently, GM3 species may function as endogenous ligands that enhance TLR4-mediated chronic inflammation, especially with very long-chain fatty acid GM3 variants(Kanoh *et al*., 2020). The shortfall in mitochondrial-acylcarnitine/acetyl-flux elevates ROS per unit of ATP and perpetuates sterile inflammatory signaling, thereby sustaining this cycle.

#### Blood-brain barrier (BBB) dysfunction

Lipid metabolism is a key factor in maintaining the normal physiological state of the BBB and in triggering its pathological damage. The metabolomics-lipidomics profile may compromise the BBB by contributing to endothelial junction dysfunction and increased inflammatory permeability(Fu *et al*., 2025). Ceramide accumulation induces endothelial barrier dysfunction(Jernigan *et al*., 2015), whereas plasmalogen depletion eliminates a crucial antioxidant reservoir that safeguards endothelial cells under ROS stress(Zoeller *et al*., 2002).

#### Neurotoxicity

The combined metabolomics-lipidomics profile may enhance neurotoxicity by linking membrane susceptibility to microglial inflammasome activation and mitochondrial energy dysfunction. The depletion of plasmalogen/ether-lipid and low levels of lysophospholipids diminish endogenous antioxidant buffering and membrane repair capacities. Zoeller et al. demonstrated that plasmalogen-deficient individuals exhibit heightened sensitivity to ROS(Zoeller *et al*., 1999). The mitochondrial-acylcarnitine/acetyl-flux deficit module indicates compromised acetyl-CoA buffering and diminished bioenergetic flexibility, which aligns with limited synaptic plasticity and lower stress resilience(Nasca *et al*., 2018). These “capacity deficits” promote diversion into the ceramide-GM3 effector module, which directly enhances neuroinflammation(Scheiblich *et al*., 2017). Significantly, the rise in ceramide is not solely correlational; Schumacher et al. established a connection between plasma ceramides and the severity of MDD, demonstrating that ceramide neutralization inhibits depressive-like behavior in mice (Schumacher *et al*., 2022). Oxidative membrane damage, mitochondrial dysfunction, and ceramide-induced microglial activation all provide plausible molecular pathways to synaptic injury, network failure, and neuroprogression.

#### MetS and atherosclerosis

The integrated architecture of 6 metabolomics and 3 lipidomics is mechanistically aligned with the advancement of metabolic syndrome and atherogenicity, as it combines deficient lipid remodeling/antioxidant buffering, mitochondrial fuel-handling inadequacy, and a sphingolipid “effector” program that disrupts and impairs insulin and endothelial signaling.

An essential athermetabolic catalyst is the increased ceramide-GM3 complex. Ceramides are recognized as mediators of lipotoxic insulin resistance and serve as predictors of cardiovascular mortality beyond LDL-C in coronary disease(Chavez & Summers, 2012; Laaksonen *et al*., 2016). Concurrently, GM3 ganglioside functions as a negative regulator of insulin signaling; the genetic ablation of GM3 synthase enhances insulin receptor phosphorylation and confers protection against diet-induced insulin resistance(Yamashita *et al*., 2003). Consequently, increased ceramides/GM3 likely contribute to insulin resistance, abnormal lipid metabolism, and endothelial dysfunction.

The two decreased lipidomic modules exacerbate atherogenic risk. The lysophospholid/ether lipid depletion module indicates a reduction in plasmalogen-mediated antioxidant protection and compromised membrane repair, and plasmalogen levels have been correlated with cardiovascular mortality(Stenvinkel *et al*., 2004), and a plasmalogen score is associated with cardiometabolic outcomes in longitudinal studies(Beyene *et al*., 2024). The mitochondrial fatty acid oxidation/acetyl-flux deficient module correlates with diminished metabolic flexibility, and these bioenergetic limitations promote lipid diversion into ceramides/DAGs.

### Lipidomics-driven Diagnostic Precision

Using PLS-derived case-wise contribution scores, we showed that, despite similar overall depression rating scores, each patient has a distinct assembly of 6 metabolic functions and 3 lipidomic modules. This implies that MDD is a spectrum of partially overlapping metabotypes and lipotypes rather than a single metabolic entity within the NIMETOX. In cases with high lipotoxicity or PL remodeling, personalized profiles can direct mechanism-based treatment decisions, favoring lipoxygenase-targeted or anti-inflammatory therapies. In cases with low Reti detox pathology, mitochondrial support is particularly important. This is consistent with nomothetic precision psychiatry, which bases therapy sequencing and selection on biological factors rather than syndromic classifications(Maes *et al*., 2022; Maes *et al*., 2024b).

The integration of metabolomic and lipidomic scores with clinical data, along with supplementary biomarkers such as inflammatory, oxidative, microbiotic, and genetic factors, may enable multimodal classification into physiologically coherent groups, rather than relying on trial-and-error therapy. Future studies may assess therapeutic algorithms that randomize patients not only on symptoms but also on metabolic and lipidomic phenotypes. For example, some patients are featured as “lipotoxic-mitochondrial” dysfunction, and other patients stand out for “inflammatory-membrane remodeling” types. These strategies would facilitate the evaluation of whether aligning drugs with the primary lipid-redox pathophysiology enhances response rates and reduces non-response related to standard therapy. Despite the results demonstrating a strong biological signal, such elevated precision scores should be approached with caution and validated in a larger, independent dataset. In the future, converting these metabotypes and lipotypes into clinically relevant decision algorithms will be crucial for advancing from descriptive lipidomics to precise personalized medicine in MDD.

## Limitations

This cross-sectional study does not allow for strong causal inference or for distinguishing between state and trait effects. Residual confounding from diet, lifestyle, and other environmental variables remains a potential risk. Future studies should investigate the link with other NIMETOX pathways, such as immune activation, oxidative stress, antioxidant defenses, and dysfunctional reverse cholesterol transport. Subsequent research should evaluate whether alterations to these pathways confer clinical benefits. External validation and longitudinal studies are crucial for assessing the accuracy of the developed lipidomic panel and pathway signatures in predicting therapy response.

## Conclusion

As this study demonstrates, serum lipidomics may identify a lipotype that precisely characterizes MDD, and the severity of phenome, which includes overall severity of disease, physiosomatic symptoms, suicidal ideation, and recurrence of illness. The featured pathways comprise the Ceramide-GM3-LNAPE (CERLNAPE) module, the mitochondrial fatty acid oxidation/acetyl-flux (CARSM) module, and the lysophospholipid/ether-lysolipid (LYSOLPE) module. Our data delineates a complex metabolic framework associated with MDD by integrating the three lipidomic modules and six metabolomics pathways. The most important features of MDD are ceramide-centered lipotoxicity and depletion of plasmalogen-carnitine pathways. These modules represent a cohesive biochemical network rather than isolated abnormalities. Based on these features, the individualized profiles can guide mechanism-based treatment choice, making personalized medicine a reality. In addition, the identified pathways highlight innovative treatment targets, such as ceramide, plasmalogens, and acetyl-carnitine. Agents that reduce ceramide, restore PL remodeling, and protect mitochondria would be beneficial.

## Supporting information

Electronic Supplementary File

## Ethical approval and consent to participate

This study obtained approval from the ethics committee of Sichuan Provincial People’s Hospital “Ethics (Research) 2024-203” and was executed in strict compliance with ethical standards and privacy regulations. All participants provided informed consent.

## Author’s contribution

Yingqian Zhang: conceptualization, sample processing, writing, and review. Xu Zhang: conceptualization, patient recruitment. Mengqi Niu: project coordination; Yiping Luo and Jing Li: patient recruitment. Abbas F. Almulla: editing, visualization; Annabel Maes: data analysis; Bo Zhou: validation and funding; Michael Maes: supervision, funding, data analysis, writing, review, and editing.

## Availability of data

The data supporting the findings of this study can be obtained upon request from the corresponding author. The data are inaccessible to the public owing to privacy or ethical constraints.

## References

Alberti, K.G., Eckel, R.H., Grundy, S.M., Zimmet, P.Z., Cleeman, J.I., Donato, K.A., Fruchart, J.C., James, W.P., Loria, C.M. & Smith, S.C., Jr. (2009) Harmonizing the metabolic syndrome: a joint interim statement of the International Diabetes Federation Task Force on Epidemiology and Prevention; National Heart, Lung, and Blood Institute; American Heart Association; World Heart Federation; International Atherosclerosis Society; and International Association for the Study of Obesity. Circulation, 120, 1640–1645.

Almulla, A.F., Thipakorn, Y., Algon, A.A.A., Tunvirachaisakul, C., Al-Hakeim, H.K. & Maes, M. (2023) Reverse cholesterol transport and lipid peroxidation biomarkers in major depression and bipolar disorder: A systematic review and meta-analysis. Brain Behav Immun, 113, 374–388.

Bazinet, R.P. & Layé, S. (2014) Polyunsaturated fatty acids and their metabolites in brain function and disease. Nat Rev Neurosci, 15, 771–785.

Beck, A.T., Steer, R.A. & Brown, G.K. (1996) BDI-II, Beck depression inventory : manual. Psychological Corp. ; Harcourt Brace San Antonio, Tex., Boston, San Antonio, Tex., Boston.

Beyene, H.B., Huynh, K., Wang, T., Paul, S., Cinel, M., Mellett, N.A., Olshansky, G., Meikle, T.G., Watts, G.F., Hung, J., Hui, J., Beilby, J., Blangero, J., Moses, E.K., Shaw, J.E., Magliano, D.J., Giles, C. & Meikle, P.J. (2024) Development and validation of a plasmalogen score as an independent modifiable marker of metabolic health: population based observational studies and a placebo-controlled cross-over study. EBioMedicine, 105, 105187.

Bigio, B., Azam, S., Mathé, A.A. & Nasca, C. (2024) The neuropsychopharmacology of acetyl-L-carnitine (LAC): basic, translational and therapeutic implications. Discov Ment Health, 4, 2.

Bisle, E., Haange, S.-B., Rojas, R., Behnke, A., Karabatsiakis, A., Gumpp, A., Mack, M., Mavioglu, R.N., Lutz-Bonengel, S., Rolle-Kampczyk, U., Mielcarek, A., von Bergen, M. & Kolassa, I.-T. (2025) Serum metabolomics in women with major depressive disorder: Associations with mitochondrial function, inflammation, and oxidative stress. Psychiatry Research, 351, 116569.

Brunkhorst-Kanaan, N., Klatt-Schreiner, K., Hackel, J., Schröter, K., Trautmann, S., Hahnefeld, L., Wicker, S., Reif, A., Thomas, D., Geisslinger, G., Kittel-Schneider, S. & Tegeder, I. (2019) Targeted lipidomics reveal derangement of ceramides in major depression and bipolar disorder. Metabolism, 95, 65–76.

Carli, F., Della Pepa, G., Sabatini, S., Vidal Puig, A. & Gastaldelli, A. (2024) Lipid metabolism in MASLD and MASH: From mechanism to the clinic. JHEP Rep, 6, 101185.

Chavez, J.A. & Summers, S.A. (2012) A ceramide-centric view of insulin resistance. Cell Metab, 15, 585–594.

Chen, T., Niu, M., Luo, Y., Li, J., Almulla, A.F., Zhang, Y. & Maes, M. (2025) The acute phase inflammatory response as a key determinant of reduced lipid-associated antioxidant defenses in Chinese patients with major depressive disorder. medRxiv, 2025.2010.2008.25337557.

Crivelli, S.M., Quadri, Z., Elsherbini, A., Vekaria, H.J., Sullivan, P.G., Zhi, W., Martinez-Martinez, P., Spassieva, S.D. & Bieberich, E. (2024) Abnormal Regulation of Mitochondrial Sphingolipids during Aging and Alzheimer’s Disease. ASN Neuro, 16, 2404367.

Dean, J.M. & Lodhi, I.J. (2017) Structural and functional roles of ether lipids. Protein Cell, 9, 196–206.

Dorninger, F., Forss-Petter, S., Wimmer, I. & Berger, J. (2020) Plasmalogens, platelet-activating factor and beyond - Ether lipids in signaling and neurodegeneration. Neurobiol Dis, 145, 105061.

Fries, G.R., Saldana, V.A., Finnstein, J. & Rein, T. (2022) Molecular pathways of major depressive disorder converge on the synapse. Mol Psychiatry, 28, 284–297.

Fu, L., Luo, T., Hao, Z., Pan, Y., Xin, W., Zhang, L., Lai, Z., Zhang, H., Liu, H. & Wei, W. (2025) Exploring novel roles of lipid droplets and lipid metabolism in regulating inflammation and blood-brain barrier function in neurological diseases. Front Neurosci, 19, 1603292.

Ge, Z., Hu, Y., Kan, W., Li, L., Xu, J., Zhang, Y., Zheng, N., Wang, G. & Du, J. (2025) Lipid metabolic dysregulation-induced neuroinflammation in the pathophysiology of major depressive disorder. Front Immunol, 16, 1625087.

Gierk, B., Kohlmann, S., Kroenke, K., Spangenberg, L., Zenger, M., Brähler, E. & Löwe, B. (2014) The somatic symptom scale-8 (SSS-8): a brief measure of somatic symptom burden. JAMA Intern Med, 174, 399–407.

Hällqvist, J., Toomey, C.E., Pinto, R., Baldwin, T., Doykov, I., Wernick, A., Al Shahrani, M., Evans, J.R., Lachica, J., Pope, S., Heales, S., Eaton, S., Mills, K., Gandhi, S. & Heywood, W.E. (2025) Multi-omic analysis reveals lipid dysregulation associated with mitochondrial dysfunction in parkinson’s disease brain. Nat Commun, 16, 10490.

Hamilton, M. (1959) The assessment of anxiety states by rating. British journal of medical psychology.

Hamilton, M. (1960) A rating scale for depression. Journal of neurology, neurosurgery, and psychiatry, 23, 56.

He, L., Duan, N., Wang, C., Shan, R., Li, J., Wang, L., Liu, Q., Tao, J., Liu, L., Ma, X. & Cao, B. (2025) Lipidomic analyses reveal the dysregulation of oxidized fatty acids (OxFAs) and acyl-carnitines (CARs) in major depressive disorder: a case-control study. BMC Psychiatry, 25, 752.

Hossain, M.I., Lee, J.H., Gagné, J.-P., Khan, J., Poirier, G.G., King, P.H., Dawson, V.L., Dawson, T.M. & Andrabi, S.A. (2024) Poly(ADP-ribose) mediates bioenergetic defects and redox imbalance in neurons following oxygen and glucose deprivation. FASEB J, 38, e23556.

Huang, C. & Chen, J.-T. (2020) Chronic retinoic acid treatment induces affective disorders by impairing the synaptic plasticity of the hippocampus. J Affect Disord, 274, 678–689.

Jansen, R., Milaneschi, Y., Schranner, D., Kastenmuller, G., Arnold, M., Han, X., Dunlop, B.W., Rush, A.J., Kaddurah-Daouk, R. & Penninx, B.W.J.H. (2024) The metabolome-wide signature of major depressive disorder. Mol Psychiatry, 29, 3722–3733.

Jernigan, P.L., Makley, A.T., Hoehn, R.S., Edwards, M.J. & Pritts, T.A. (2015) The role of sphingolipids in endothelial barrier function. Biol Chem, 396, 681–691.

Jirakran, K., Almulla, A.F., Jaipinta, T., Vasupanrajit, A., Jansem, P., Tunvirachaisakul, C., Dzhambazova, E., Stoyanov, D.S. & Maes, M. (2025a) Increased atherogenicity in mood disorders: a systematic review, meta-analysis and meta-regression. Neurosci Biobehav Rev, 169, 106005.

Jirakran, K., Vasupanrajit, A., Tunvirachaisakul, C., Almulla, A.F., Kubera, M. & Maes, M. (2025b) Lipid profiles in major depression, both with and without metabolic syndrome: associations with suicidal behaviors and neuroticism. BMC Psychiatry, 25, 379.

Kanoh, H., Nitta, T., Go, S., Inamori, K.-I., Veillon, L., Nihei, W., Fujii, M., Kabayama, K., Shimoyama, A., Fukase, K., Ohto, U., Shimizu, T., Watanabe, T., Shindo, H., Aoki, S., Sato, K., Nagasaki, M., Yatomi, Y., Komura, N., Ando, H., Ishida, H., Kiso, M., Natori, Y., Yoshimura, Y., Zonca, A., Cattaneo, A., Letizia, M., Ciampa, M., Mauri, L., Prinetti, A., Sonnino, S., Suzuki, A. & Inokuchi, J.-I. (2020) Homeostatic and pathogenic roles of GM3 ganglioside molecular species in TLR4 signaling in obesity. EMBO J, 39, e101732.

Knowles, E.E.M., Huynh, K., Meikle, P.J., Göring, H.H.H., Olvera, R.L., Mathias, S.R., Duggirala, R., Almasy, L., Blangero, J., Curran, J.E. & Glahn, D.C. (2017) The lipidome in major depressive disorder: Shared genetic influence for ether-phosphatidylcholines, a plasma-based phenotype related to inflammation, and disease risk. Eur Psychiatry, 43, 44–50.

Laaksonen, R., Ekroos, K., Sysi-Aho, M., Hilvo, M., Vihervaara, T., Kauhanen, D., Suoniemi, M., Hurme, R., März, W., Scharnagl, H., Stojakovic, T., Vlachopoulou, E., Lokki, M.-L., Nieminen, M.S., Klingenberg, R., Matter, C.M., Hornemann, T., Jüni, P., Rodondi, N., Räber, L., Windecker, S., Gencer, B., Pedersen, E.R., Tell, G.S., Nygård, O., Mach, F., Sinisalo, J. & Lüscher, T.F. (2016) Plasma ceramides predict cardiovascular death in patients with stable coronary artery disease and acute coronary syndromes beyond LDL-cholesterol. Eur Heart J, 37, 1967–1976.

Liu, J., Fan, X., Song, Y. & Zhao, J. (2025) Triglyceride-based lipotoxicity in the pathophysiology of chronic diseases. Trends Endocrinol Metab, S1043-2760, 00150-X.

Maes, M., Almulla, A.F., Stoyanov, D. & Zhang, Y. (2025a) Advancements in the molecular understanding of major depressive disorder uncovering novel targets with therapeutic promise: focus on recurrence of illness. Expert Opin Ther Targets.

Maes, M., Almulla, A.F., You, Z. & Zhang, Y. (2025b) Neuroimmune, metabolic and oxidative stress pathways in major depressive disorder. Nat Rev Neurol, Sep; 21, 473–489.

Maes, M., Delanghe, J., Meltzer, H.Y., Scharpé, S., D’Hondt, P. & Cosyns, P. (1994) Lower degree of esterification of serum cholesterol in depression: relevance for depression and suicide research. Acta Psychiatr Scand, 90, 252–258.

Maes, M., Jirakran, K., Semeão, L.d.O., Michelin, A.P., Matsumoto, A.K., Brinholi, F.F., Barbosa, D.S., Tivirachaisakul, C., Almulla, A.F., Stoyanov, D. & Zhang, Y. (2025c) Key factors underpinning neuroimmune-metabolic-oxidative (NIMETOX) major depression in outpatients: paraoxonase 1 activity, reverse cholesterol transport, increased atherogenicity, protein oxidation, and differently expressed cytokine networks. Neuro Endocrinol Lett, 46, 115–125.

Maes, M., Jirakran, K., Vasupanrajit, A., Niu, M., Zhou, B., Stoyanov, D.S. & Tunvirachaisakul, C. (2024a) The recurrence of illness (ROI) index is a key factor in major depression that indicates increasing immune-linked neurotoxicity and vulnerability to suicidal behaviors. Psychiatry Res, 339, 116085.

Maes, M., Moraes, J.B., Congio, A., Vargas, H. & Nunes, S. (2022) Research and Diagnostic Algorithmic Rules (RADAR) for mood disorders, recurrence of illness, suicidal behaviours, and the patient’s lifetime trajectory. Acta Neuropsychiatr, 35, 104–117.

Maes, M., Niu, M., Maes, A., Luo, Y., Yangyang, C., Li, J., Almulla, A.F. & Zhang, Y. (2025d) Peripheral Metabolic-Redox Signaling as a Core Mechanism of Major Depressive Disorder: Evidence From Deep Metabolomic Phenotyping. medRxiv, 2025.2012.2015.25342323.

Maes, M., Niu, M., Wang, P., Maes, A., Luo, Y., Yangyang, C., Zhuang, X., Almulla, A.F., Li, J. & Zhang, Y. (2026) NIMETOX-informed Precision Nomothetic Models of Major Depressive Disorder: Group, Phenome, and Individual Signatures. medRxiv, 2026.2001.2023.26344678.

Maes, M., Smith, R., Christophe, A., Vandoolaeghe, E., Van Gastel, A., Neels, H., Demedts, P., Wauters, A. & Meltzer, H.Y. (1997) Lower serum high-density lipoprotein cholesterol (HDL-C) in major depression and in depressed men with serious suicidal attempts: relationship with immune-inflammatory markers. Acta Psychiatr Scand, 95, 212–221.

Maes, M., Zhou, B., Jirakran, K., Vasupanrajit, A., Boonchaya-Anant, P., Tunvirachaisakul, C., Tang, X., Li, J. & Almulla, A.F. (2024b) Towards a major methodological shift in depression research by assessing continuous scores of recurrence of illness, lifetime and current suicidal behaviors and phenome features. J Affect Disord, 350, 728–740.

Malhi, G.S. & Mann, J.J. (2018) Depression. Lancet, 392, 2299–2312.

Miao, G., Deen, J., Struzeski, J.B., Chen, M., Zhang, Y., Cole, S.A., Fretts, A.M., Lee, E.T., Howard, B.V., Fiehn, O. & Zhao, J. (2023) Plasma lipidomic profile of depressive symptoms: a longitudinal study in a large sample of community-dwelling American Indians in the strong heart study. Mol Psychiatry, 28, 2480–2489.

Montanari, S., Jansen, R., Schranner, D., Kastenmüller, G., Arnold, M., Janiri, D., Sani, G., Bhattacharyya, S., Mahmoudian Dehkordi, S., Dunlop, B.W., Rush, A.J., Penninx, B.W.H.J., Kaddurah-Daouk, R. & Milaneschi, Y. (2025) Acylcarnitines metabolism in depression: association with diagnostic status, depression severity and symptom profile in the NESDA cohort. Transl Psychiatry, 15, 65.

Nasca, C., Bigio, B., Lee, F.S., Young, S.P., Kautz, M.M., Albright, A., Beasley, J., Millington, D.S., Mathé, A.A., Kocsis, J.H., Murrough, J.W., McEwen, B.S. & Rasgon, N. (2018) Acetyl-l-carnitine deficiency in patients with major depressive disorder. Proc Natl Acad Sci U S A, 115, 8627–8632.

Niu, M., Zhang, Y., Zhang, X., Yangyang, C., Zhuang, X., Li, J. & Maes, M. (2025) Dissection of the clinical phenome of major depressive disorder into subdomains and subgroups in relation to activated immune-inflammatory profiles. medRxiv, 2025.2010.2021.25338439.

Posner, K., Brown, G.K., Stanley, B., Brent, D.A., Yershova, K.V., Oquendo, M.A., Currier, G.W., Melvin, G.A., Greenhill, L., Shen, S. & Mann, J.J. (2011) The Columbia-Suicide Severity Rating Scale: initial validity and internal consistency findings from three multisite studies with adolescents and adults. Am J Psychiatry, 168, 1266–1277.

Post, R.M. (2018) Myriad of implications of acetyl-l-carnitine deficits in depression. Proc Natl Acad Sci U S A, 115, 8475–8477.

Scheiblich, H., Schlütter, A., Golenbock, D.T., Latz, E., Martinez-Martinez, P. & Heneka, M.T. (2017) Activation of the NLRP3 inflammasome in microglia: the role of ceramide. J Neurochem, 143, 534–550.

Schumacher, F., Edwards, M.J., Mühle, C., Carpinteiro, A., Wilson, G.C., Wilker, B., Soddemann, M., Keitsch, S., Scherbaum, N., Müller, B.W., Lang, U.E., Linnemann, C., Kleuser, B., Müller, C.P., Kornhuber, J. & Gulbins, E. (2022) Ceramide levels in blood plasma correlate with major depressive disorder severity and its neutralization abrogates depressive behavior in mice. J Biol Chem, 298, 102185.

Serbulea, V., DeWeese, D. & Leitinger, N. (2017) The effect of oxidized phospholipids on phenotypic polarization and function of macrophages. Free Radic Biol Med, 111, 156–168.

Shmarakov, I.O. (2015) Retinoid-xenobiotic interactions: the Ying and the Yang. Hepatobiliary Surg Nutr, 4, 243–267.

Song, Y., Cao, H., Zuo, C., Gu, Z., Huang, Y., Miao, J., Fu, Y., Guo, Y., Jiang, Y. & Wang, F. (2023) Mitochondrial dysfunction: A fatal blow in depression. Biomed Pharmacother, 167, 115652.

Spielberger, C., Gorsuch, R., Lushene, R., Vagg, P. & Jacobs, G. (1983) Manual for the State-Trait Anxiety Inventory; Palo Alto, CA, Ed. Palo Alto: Spielberger.

Stenvinkel, P., Diczfalusy, U., Lindholm, B. & Heimbürger, O. (2004) Phospholipid plasmalogen, a surrogate marker of oxidative stress, is associated with increased cardiovascular mortality in patients on renal replacement therapy. Nephrol Dial Transplant, 19, 972–976.

Tkachev, A., Stekolshchikova, E., Golubova, A., Serkina, A., Morozova, A., Zorkina, Y., Riabinina, D., Golubeva, E., Ochneva, A., Savenkova, V., Petrova, D., Andreyuk, D., Goncharova, A., Alekseenko, I., Kostyuk, G. & Khaitovich, P. (2024) Screening for depression in the general population through lipid biomarkers. EBioMedicine, 110, 105455.

van der Spek, A., Stewart, I.D., Kühnel, B., Pietzner, M., Alshehri, T., Gauß, F., Hysi, P.G., MahmoudianDehkordi, S., Heinken, A., Luik, A.I., Ladwig, K.-H., Kastenmüller, G., Menni, C., Hertel, J., Ikram, M.A., de Mutsert, R., Suhre, K., Gieger, C., Strauch, K., Völzke, H., Meitinger, T., Mangino, M., Flaquer, A., Waldenberger, M., Peters, A., Thiele, I., Kaddurah-Daouk, R., Dunlop, B.W., Rosendaal, F.R., Wareham, N.J., Spector, T.D., Kunze, S., Grabe, H.J., Mook-Kanamori, D.O., Langenberg, C., van Duijn, C.M. & Amin, N. (2023) Circulating metabolites modulated by diet are associated with depression. Mol Psychiatry, 28, 3874–3887.

Vanherle, S., Loix, M., Miron, V.E., Hendriks, J.J.A. & Bogie, J.F.J. (2025) Lipid metabolism, remodelling and intercellular transfer in the CNS. Nat Rev Neurosci, 26, 214–231.

Wang, B. & Tontonoz, P. (2019) Phospholipid Remodeling in Physiology and Disease. Annual Review of Physiology, 81, 165–188.

Wang, J., Hu, X., Li, Y., Li, S., Wang, T., Wang, D., Gao, Y., Wang, Q., Zhou, J. & Wan, C. (2025) Impaired lipid homeostasis and elevated lipid oxidation of erythrocyte membrane in adolescent depression. Redox Biol, 80, 103491.

Wilkerson, J.L., Tatum, S.M., Holland, W.L. & Summers, S.A. (2024) Ceramides are fuel gauges on the drive to cardiometabolic disease. Physiol Rev, 104, 1061–1119.

Xiang, F., Zhang, Z., Xie, J., Xiong, S., Yang, C., Liao, D., Xia, B. & Lin, L. (2025) Comprehensive review of the expanding roles of the carnitine pool in metabolic physiology: beyond fatty acid oxidation. J Transl Med, 23, 324.

Yamashita, T., Hashiramoto, A., Haluzik, M., Mizukami, H., Beck, S., Norton, A., Kono, M., Tsuji, S., Daniotti, J.L., Werth, N., Sandhoff, R., Sandhoff, K. & Proia, R.L. (2003) Enhanced insulin sensitivity in mice lacking ganglioside GM3. Proc Natl Acad Sci U S A, 100, 3445–3449.

Zachrisson, O., Regland, B., Jahreskog, M., Kron, M. & Gottfries, C.G. (2002) A rating scale for fibromyalgia and chronic fatigue syndrome (the FibroFatigue scale). J Psychosom Res, 52, 501–509.

Zoeller, R.A., Grazia, T.J., LaCamera, P., Park, J., Gaposchkin, D.P. & Farber, H.W. (2002) Increasing plasmalogen levels protects human endothelial cells during hypoxia. Am J Physiol Heart Circ Physiol, 283, H671–H679.

Zoeller, R.A., Lake, A.C., Nagan, N., Gaposchkin, D.P., Legner, M.A. & Lieberthal, W. (1999) Plasmalogens as endogenous antioxidants: somatic cell mutants reveal the importance of the vinyl ether. Biochem J, 338 ( **Pt 3**), 769–776.

