## Supplementary material for "Deep Lipidomic Phenotyping Identifies Ceramide-Centered Lipotoxicity and Depletion of Plasmalogen-Carnitine Pathways in Major Depressive Disorder: Implications for Precision Medicine": Electronic Supplementary File

Electronic Supplementary File (ESF)

Supplementary methods:

*Assay*

Metabolites extract: A 200 μL sample was combined with 400 μL of cold Methyl tert-Butyl Ether (MTBE) solution and 80 μL of icy methanol. Centrifuge at 3000 rpm for 15 minutes and transfer 200 μL of the supernatant to a separate EP tube. Following the freeze-drying of the supernatant, 200 μL of a dichloromethane and methanol mixture (v/v = 1:1) was introduced for reconstitution. Subsequently, centrifugation was performed at 3000 rpm for 15 minutes, after which the supernatant was transferred to a new vial for UPLC-HRMS analysis. The quality control (QC) sample was formulated by combining equal amounts of the supernatants from the samples.

Chromatography: An ACQUITY UPLC CSH C18 column (100 mm × 2.1 mm, 1.7 µm, Waters) was employed for separation. The mobile phase comprises phase A (Acetonitrile: water (6:4) + 10 mmol/L ammonium formate + 0.1% formic acid) and phase B (Isopropanol: acetonitrile (9:1) + 10 mmol/L ammonium formate + 0.1% formic acid). The gradient elution parameters were established as follows: 0 to 0.4 minutes at 30% B; 0.4 to 1.0 minutes from 30% to 45% B; 1.0 to 3.5 minutes from 45% to 60% B; 3.5 to 5.0 minutes from 60% to 75% B; 5.0 to 7.0 minutes from 75% to 90% B; 7.0 to 8.5 minutes at 90% to 100% B; 8.5 to 8.6 minutes at 100% B; 8.6 to 8.61 minutes from 100% to 30% B; 8.61 to 10.0 minutes at 30% B. The flow rate is 0.3 mL/min. The injection volume for each specimen was 4 µL. The column oven was regulated at 40℃.

Mass spectrometry: A high-resolution tandem mass spectrometer, the Triple TOF 6600 (AB SCIEX), was employed to detect lipids eluted from the column. Each sample was analyzed using both positive and negative electrospray ionization modes. The ESI temperature is 500 degrees Celsius. The voltage is +5000 volts in positive ion mode and -4500 volts in negative ion mode. The curtain gas pressure of the ion source is 30 psi, while the pressures for Gas 1 (Auxiliary gas) and Gas 2 (Sheath gas) are both set at 60 psi. The mass spectrometric data were acquired using full scan and information-dependent acquisition (IDA) modes. During a single acquisition cycle, the complete scan range is 50-2000 Da, with a full-scan acquisition duration of 170 ms. The top 12 signal ions, with signal accumulation intensity>100, were selected from the full scan for IDA analysis, with an acquisition range of 25-1200 Da and an acquisition duration of 30 ms. The dynamic exclusion is established at 4 seconds.

*LC-MS raw data processing*

The MS data preprocessing, which encompassed peak picking, peak grouping, retention time correction, secondary peak grouping, and annotation of isotopes and adducts, was executed utilizing XCMS software. LC-MS raw data files were converted to mzXML format and subsequently analyzed using the XCMS, CAMERA, and metaX toolboxes in R. Each ion was characterized by integrating retention time (RT) with m/z data. The intensities of each peak were documented, resulting in a three-dimensional matrix of arbitrarily designated peak indices (retention time-m/z pairings), sample names (observations), and ion intensity data (variables).

The online KEGG and HMDB databases were used to annotate the metabolites by correlating the precise molecular mass (m/z) data of the samples with those in the databases. If the mass discrepancy between the observed and database values was less than 10 ppm, the metabolite would be annotated, and its chemical formula would be determined and validated using isotopic distribution measurements. We also employed an internal fragment-spectrum library of metabolites to corroborate metabolite identifications.

*Machine learning for biomarker discovery*

PCA with subsequent varimax rotation was employed to identify latent constructs from the lipidomic dataset. The Kaiser-Meyer-Olkin (KMO) measure was employed to assess sampling adequacy, with values above 0.7 indicating sufficient factorability. Principal components were retained if they explained more than 50% of the variance and demonstrated factor loadings greater than 0.6 on all contributing variables.

Centered Log-Ratio (CLR) transformations of the metabolic data were implemented in R due to the compositional nature of lipidomics data, which is constrained by constant-sum scaling, making it vulnerable to spurious correlations and misleading patterns when evaluated on raw or non-ratio-based scales. Comprehensive, curated, and selected CLR-transformed datasets were analyzed to determine the multivariate difference between MDD and HC using all or curated lipids. We started with endogenous and external lipids data (n = 1095) from the comprehensive lipids matrix. The entire curated lipidomics matrix of endogenous lipids was then examined. Next, statistical analysis was predominantly performed on selected lipids using R (v4.0). Metabolite data underwent three primary processing steps: first, data filtering to exclude samples with over 80% missing values or quality control (QC) samples with over 50% missing data; second, data imputation utilizing the K-nearest neighbor (KNN) method; and third, data standardization through Probabilistic Quotient Normalization (PQN). Cluster heatmaps were produced using the R package pheatmap. PCA and significant differential metabolite analysis were conducted using the R package ropls, and variable importance in projection (VIP) values were computed for each variable. Correlation analysis was performed utilizing Pearson's correlation coefficient from the R package cor. The conclusive differential metabolites were determined using three criteria: *P*-value < 0.05 from t-test, fold change > 1.2, and VIP ≥ 1 from PLS-DA analysis. The PLS-DA model was evaluated using R^2^Y (explained variance in group membership), Q^2^ (cross-validated predictive performance), and CV-ANOVA on 1095 and 157 lipids to determine the statistical significance of class separation. VIP scores were used to identify the variables most affecting this multivariate structure, and the investigation was repeated using the highest-ranked VIP features to demonstrate their combined impact on PLS-DA discrimination. PLS-DA on the whole dataset (exogenous + endogenous) and VIP-selected metabolites from the curated dataset will be shown. This method was used just for exploratory multivariate analysis, not biomarker identification, predictive modeling, or validation.

ESF, Table 1. Demographic and clinical data of MDD patients and healthy controls

| **Variables** | **HC (n = 40)** | **MDD (n = 125)** | **F/χ^2^** | **df** | ***p*** |
| --- | --- | --- | --- | --- | --- |
| Age (years) | 37.1 (13.8) | 35.7 (12.1) | 0.37 | 1/163 | 0.542 |
| Sex (Female/Male) | 27/13 | 87/37 | 0.06 | 1 | 0.802 |
| Marriage status (Yes/No) | 17/23 | 69/56 | 1.96 | 1 | 0.162 |
| Living conditions (Urban/Rural) | 36/4 | 114/11 | FET | - | 0.760 |
| Educational years | 13.9 (4.3) | 13.5 (3.3) | 0.26 | 1/163 | 0.613 |
| BMI (kg/m^2^) | 23.52 (4.07) | 22.30 (3.36) | 3.58 | 1/163 | 0.06 |
| Smoking (Yes/No) | 3/37 | 24/101 | FET | - | 0.091 |
| Physiosomatic symptoms (z-score) | -1.300 (0.421) | 0.477 (0.671) | MWUT | - | < 0.001 |
| Current suicidal ideation (z-score) | -0.774 (0.262) | 0.269 (1.021) | MWUT | - | < 0.001 |
| Recurrence of illness (ROI) (z-score) | -1.057 (0.150) | 0.367 (0.903) | MWUT | - | < 0.001 |
| Overall severity of depression (OSOD) (z-score) | -1.464 (0.671) | 0.537 (0.510) | MWUT | - | < 0.001 |

Results are shown as mean (SD); F: results of analysis of variance; χ^2^: analysis of contingency table; FET: Fisher’s exact probability test; MWUT: Mann-Whitney U test.

ESF, Table 2. The lipidomic modules examined in the present study

| **Functional metabolic modules** | **Annotations (and sign in MDD)** |
| --- | --- |
| **Lipotoxicity** | ↑ 1-Stearoyl-2-arachidonoylglycerol |
| **Lipotoxicity** | ↓ DG(18:1(11Z)/18:2(9Z,12Z)/0:0) |
| **Lipotoxicity** | ↑ Myristic acid, tetradecanoic acid, C14:0 |
| **Changes in FA storage/ metabolism/signaling** | ↓ Arachidic acid, eicosanoic acid, C20:0 |
| **Changes in FA storage/ metabolism/signaling** | ↑ Methyl stearate, methyl C18:0, SAME |
| **Ether lipids** | ↓ 1-O-Hexadecyl-lyso-sn-glycero-3-phosphocholine |
| **PL remodeling** | ↑ PI(22:4(7Z,10Z,13Z,16Z)/16:0): |
| **PL remodeling** | ↑ 14,15-Leukotriene C4(ExC4), Eoxin C4 |
| **PL remodeling** | ↑ PS(22:5(4Z,7Z,10Z,13Z,16Z)/22:4(7Z,10Z,13Z,16Z)) |
| **PL remodeling** | ↑ 1-Stearoyl-2-arachidonyl-sn-glycero-3-phosphocholine |
| **PL remodeling** | ↑ PS(22:5(7Z,10Z,13Z,16Z,19Z)/22:4(7Z,10Z,13Z,16Z)) |
| **PL remodeling** | ↑ 1,2-Dioleoyl-sn-glycero-3-phospho-(1'-myo-inositol) |
| **Retinoid detox, Vit A metabolism** | ↓ rac-4-Hydroxy-4-O-(beta-D-glucuronide)-all-trans-retinyl acetate |
| **Mito ATP Redox** | ↑ Cysteylhistidine |
| **Mito ATP Redox** | ↑ Inosinic acid |
| **Mito ATP Redox** | ↑ Hydroxyphenyllactic acid (HPLA) |

FA: fatty acid

ESF, Figure 1

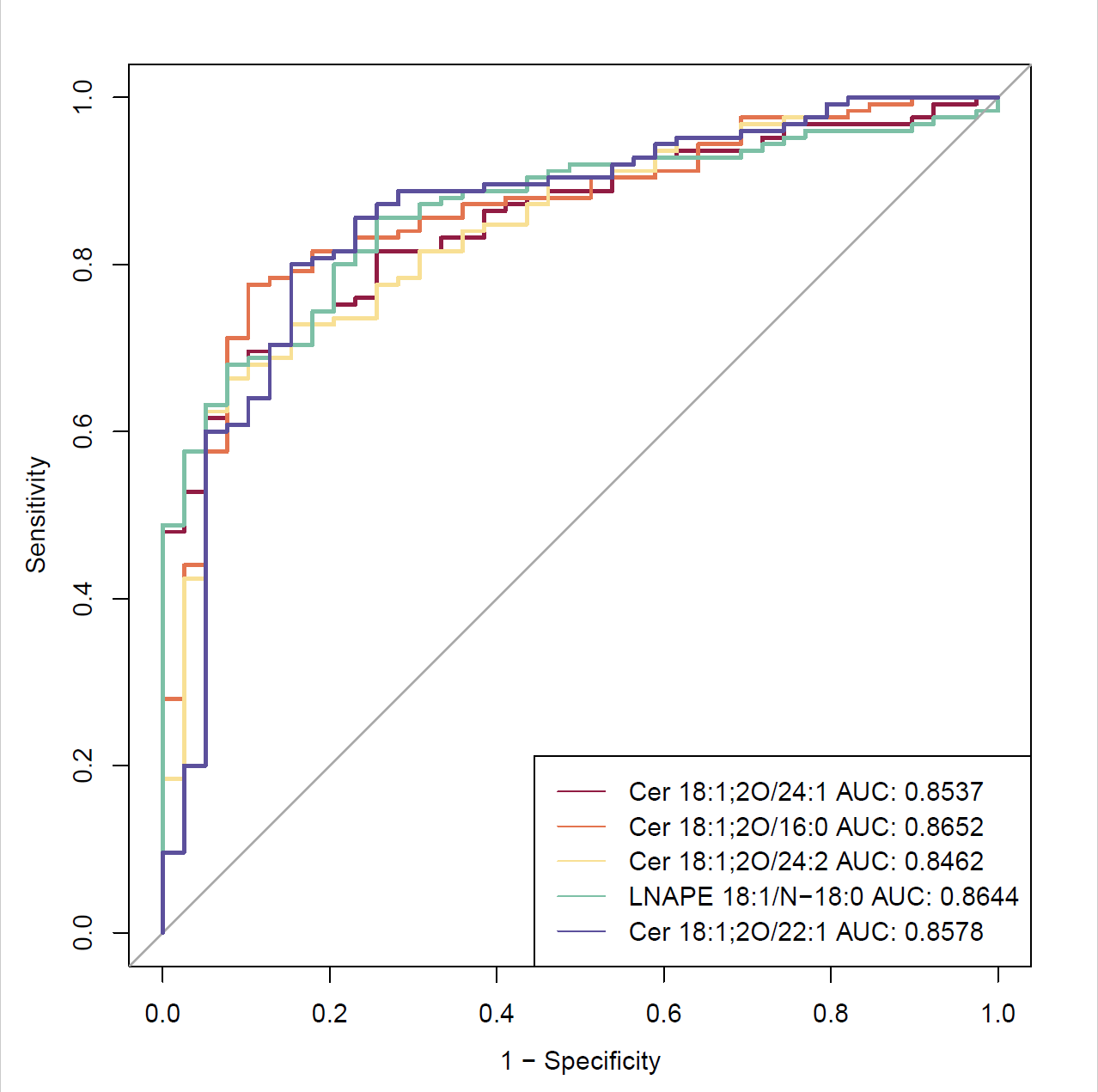

ESF, Figure 1. The ROC curve of the top 5 lipids
